# NXT2 is the key player for nuclear RNA export in the human testis and critical for spermatogenesis

**DOI:** 10.1101/2024.08.01.24310552

**Authors:** Ann-Kristin Dicke, Ammar Ahmedani, Lin Ma, Godfried W. van der Heijden, Sophie A. Koser, Claudia Krallmann, Oguzhan Kalyon, Miguel J. Xavier, Joris A. Veltman, Sabine Kliesch, Nina Neuhaus, Noora Kotaja, Frank Tüttelmann, Birgit Stallmeyer

**Affiliations:** Centre of Medical Genetics, Institute of Reproductive Genetics, University of Münster, Münster, Germany; Institute of Biomedicine, Integrative Physiology and Pharmacology Unit, University of Turku, Turku, Finland; Division of Reproductive Medicine, Department of Obstetrics and Gynecology, Radboudumc, Nijmegen, Netherlands; Centre of Reproductive Medicine and Andrology, Department of Clinical and Surgical Andrology, University Hospital Münster, Münster, Germany; Biosciences Institute, Faculty of Medical Sciences, Newcastle University, Newcastle-upon-Tyne, UK

**Keywords:** nuclear RNA export, nuclear transport, male infertility, non-obstructive azoospermia, X chromosome, NXT2, NXF2, NXF3

## Abstract

In eukaryotes, the nucleocytoplasmic export of bulk poly(A)^+^-mRNAs through the nuclear pore complex is mediated by the ubiquitously expressed NXT1-NXF1 heterodimer. In humans, *NXT1* has an X-chromosomal paralog, *NXT2,* which exhibits testis-enriched expression, suggesting a role in spermatogenesis. Here, we report the *in vivo* interaction of NXT2 with crucial components of the nuclear export machinery, including NXF1, the testis-specific NXF1 paralogs NXF2 and NXF3, and the nuclear pore complex proteins NUP93 and NUP214. Further, NXT2’s NTF2-like domain mediates binding to NXF2 and NXF3. By identifying infertile men with loss-of-function variants in *NXT2* and *NXF3*, we link the impaired NXT2-NXF activity to disturbed germ cell development. The predominant absence of germ cells in men with NXT2 deficiency indicates its critical function already during fetal or first steps of germ cell development. In contrast, loss of NXF3 affects later stages of spermatogenesis resulting in quantitatively and qualitatively impaired sperm production.

## Introduction

In eukaryotic cells, the nuclear envelope is an essential barrier with multiple functions. It separates the nucleus, where precursor messenger RNA (pre-mRNA) is transcribed and processed into mature mRNA, from the cytoplasm, where the mRNA is translated into protein. This separation allows quality control by eliminating non-functional RNAs in the nucleus before they enter the translation machinery and is, thus, involved in the regulation of gene expression^1^. In addition, the nuclear envelope prevents macromolecules from translocating freely between the two compartments and has an important structural role in sheltering the genome^2^. To regulate the nucleocytoplasmic trafficking of macromolecules, such as proteins and RNA, massive (∼120 MDa) nuclear pore complexes (NPCs), consisting of multiple copies of about 30 different nuclear pore proteins (nucleoporins/NUPs), are embedded in the nuclear envelope, forming specialized channels^3^. Transcripts that passed nuclear quality control are bound to ribonucleoproteins (RNPs), forming compact globules that are recognized and coated by transcription export (TREX) complexes, which license the handoff to the nuclear export factor heterodimer, composed of NXT1 (also known as p15/p15-1) and NXF1 (also known as TAP)^4,5^.

NXF1 is a modular protein consisting of four highly conserved domains: a nuclear localization signal, a non-canonical RNA-recognition motif domain (RRMD) essential for binding the cargo mRNA, a nuclear transport factor 2 (NTF2)-like domain that mediates binding to NXT1, and a C-terminal nuclear pore complex binding domain establishing the interaction with NUPs^6–8^. NXT1 consists almost exclusively of the NTF2-like domain and functions as a critical cofactor enhancing nuclear pore complex binding of NXF1^8^.

NXF1 is highly evolutionarily conserved and its essential role in the nuclear export of polyadenylated mRNAs is well established from yeast to mammals^9^. In humans, NXT1 and NXF1 are ubiquitously expressed and the NXT1-NXF1 export pathway is involved in bulk mRNA export in diverse tissues^10^. Interestingly, humans have X-chromosomal paralogs of *NXT1* and *NXF1*, known as *NXT2* and *NXF2*, *NXF3*, *NXF4*, and *NXF5*, which have a tissue-enriched/specific expression profile. NXT2 is specific to eutherians and shares ∼75 % amino acid sequence similarity with NXT1^7^. It binds to NXF proteins *in vitro*^7^, but the cellular function remains unclear.

Interestingly, in eutherians, NXT2 has been demonstrated to have different expression profiles in different lineages. While mouse *Nxt2* is ubiquitously expressed, human *NXT2* shows testis-enriched expression, suggesting an evolutionarily young and specific role of the human NXT2 in spermatogenesis. Indeed, *Nxt2* has evolved conservatively in mice, implying functional constraints as predominant force^11^. In contrast, adaptive selection, which describes the propagation of advantageous genetic variations through positive selection, has frequently contributed to genetic changes in primate *NXT2*^11^. This quite different evolutionary development of the two NXT2 orthologs indicates that the primate protein has acquired novel substrate- and/or tissue-specific functions that not only differ from the function of NXT1 but also from the function of the murine NXT2^11^.

Here, we demonstrate that not only NXF1 but also its testis-specific paralogs NXF2 and NXF3 belong to the human testicular interactome of NXT2 and propose an adapted nuclear RNA export pathway in the human testis with NXT2 as the key player. In addition, we provide evidence that an impaired testis-specific NXT2 interactome leads to male infertility. Men with loss-of-function (LoF) variants in *NXT2* show absence of sperm in the ejaculate (azoospermia) and near absence of germ cells in their testes (non-obstructive azoospermia, NOA). In contrast, a man with a LoF in *NXF3* produced very few, and predominantly immotile sperm. Taken together, this indicates that the NXT2 interactome likely plays critical roles during embryonic/fetal germ cell development as well as during spermatogenesis in the adult testis.

## Results

### A nuclear export pathway involving NXT2 in the adult human testis

As *NXT2* shows a testis-enriched expression in humans and has been influenced by adaptive selection in primates^11^, it is likely that the encoded protein has evolved to acquire functions and binding partners distinct from its paralog NXT1. Therefore, we aimed to identify the interactome of NXT2 *in vivo* by pulling down NXT2 from adult human testis tissue lysates using a validated antibody (Supplementary Figure 1a/b, Supplementary Table 1), followed by a mass spectrometry analysis. Eight proteins, including NXT2, were significantly enriched compared to the IgG pulldown control (Figure 1a, Supplementary Table 2). STRING interaction analysis of the pulled-down proteins highlighted three major clusters (Figure 1b). The core cluster includes NXT2 and three members of the nuclear export factor protein family, NXF1, NXF2, and NXF3. A possible function of NXT2 in RNA export through the nuclear pore was strengthened by the identification of two nucleoporins, NUP93 and NUP214, in its interactome. In addition, the mass spectrometry data indicated an interaction of NXT2 with two ribosome biogenesis factors, SPATA5 and SPATA5L1. Gene Ontology analysis further supported a role for NXT2 and its interaction partners in nucleocytoplasmic RNA transport as evidenced by the enrichment in terms mRNA transport, nuclear export and nuclear transport (Figure 1c). The main cellular component associated with the NXT2 interactome was the nuclear export factor complex (Figure 1d).

**Figure 1:**
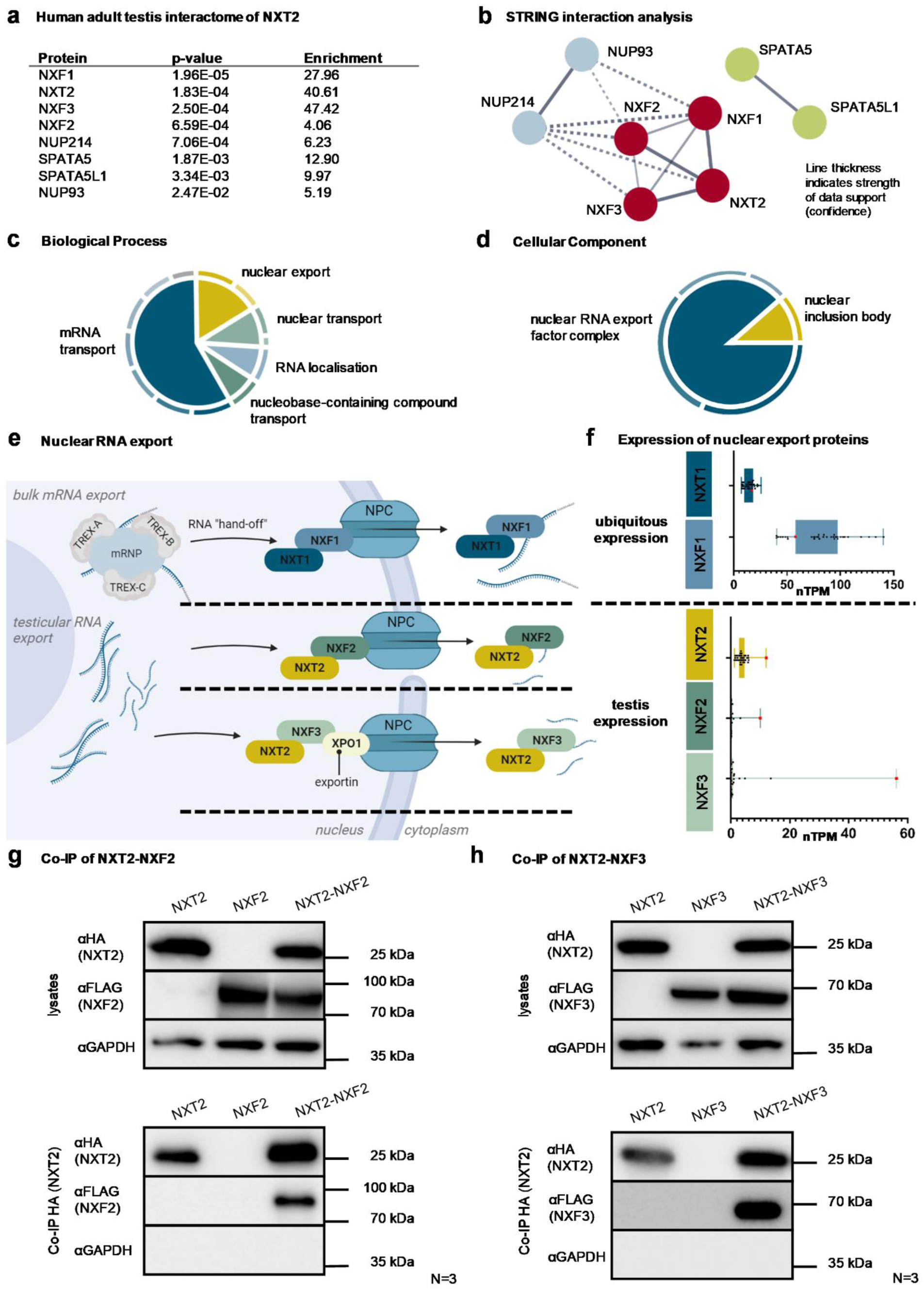
The testicular interactome of NXT2 indicates a testis-specific nucleocytoplasmic RNA export pathway. a. Results of mass spectrometry data of an NXT2 pulldown (N=4) from adult human testis depicting proteins significantly (p<0.05) and at least 4-fold enriched compared to the IgG control (N=2) (two-sided t-test on protein abundance values). b. The STRING interaction analysis of significantly enriched proteins identified three main clusters. The central cluster (red) includes NXT2 and the nuclear export factors NXF1, NXF2, and NXF3. The blue cluster comprises two nucleoporins (NUP93 and NUP214), and the green cluster includes the ribosome biogenesis factors SPATA5 and SPATA5L1. c. Gene Ontology (GO) term analysis of significantly enriched proteins highlights biological processes associated with NXT2 and its binding partners in a two-tiered-hierarchy. Top GO terms were “mRNA transport” and “nuclear export”. Terms belonging to the inner circular segment are listed. d. Name and relative proportion of GO terms describing the cellular compartment associated with the NXT2 interactome. GO terms belonging to the inner ring are indicated. e. Proposed models for testis-specific RNA export pathways with NXT2 as the key player. At the top, the well-studied ubiquitous nuclear bulk mRNA export pathway involving NXT1 and NXF1 is depicted. In the middle and at the bottom, two possible testis-specific nuclear export pathways are shown involving NXT2 and its testicular binding partners NXF2 and NXF3, respectively. f. Box plots visualizing mRNA expression data of human nuclear export factor genes derived from the Human Protein Atlas (nTPM = normalized transcripts per million). g. Western blot analysis of Co-IP of NXT2-HA and NXF2-FLAG demonstrates that NXF2 binds to NXT2 *in vitro* in lysates derived from NXT2-NXF2 co-transfection in HEK293T cells. h. Binding of NXT2 to NXF3 was corroborated in NXT2 Co-IP of overexpressed proteins as shown in subsequent Western blot analysis (bottom). Representative Western blots from at least three replicates (N = 3) are shown. Uncropped images of all Western blots depicted are provided in the Source data file.

Interestingly, NXF1 was also identified in the NXT2 interactome. In a complex with NXT1, NXF1 is well-known to be involved in the ubiquitous nuclear export of bulk mRNA via translocation through the nuclear pore complex^12^ (Figure 1e). However, NXT1 was absent from the NXT2-NXF interactome, indicating that it cannot compensate for the testicular function of NXT2 and, *vice versa*, that NXT2 is the main interaction partner of NXF1 in the human testis.

The protein structures of NXF2 and NXF3 are very similar to that of NXF1 (Supplementary Figure 2). However, in contrast to NXF1, NXF2 and 3 are encoded by X-chromosomal genes and are specifically expressed in the testis (Figure 1f), arguing for a testis-specific RNA export pathway, functionally similar to the NXT1-NXF1-mediated ubiquitous bulk mRNA export pathway, but involving testis-enriched NXT2 as the key player and NXF2 and NXF3 as testis-specific interaction partners (Figure 1e).

To corroborate the proposed direct interaction of NXT2 with NXF2 and NXF3, we performed co-immunoprecipitation (Co-IP) of HA-tagged NXT2 with FLAG-tagged NXF2 (Figure 1g) or FLAG-tagged NXF3 (Figure 1h) overexpressed in HEK293T cells. NXF2 and NFX3 were only detectable in Western blots when NXT2 was co-expressed, demonstrating a specific and direct interaction of both proteins with NXT2 also *in vitro*.

### NXT2 interacts with NXF2 and NXF3 through NTF2-like domains

NXF1, NXF2, and NXF3 share very similar protein structures, consisting of an RNA recognition motif domain (RRM domain) and a nuclear transport factor 2-like domain (NTF2-like domain/NTF2LD). NXF2 also shares a C-terminal nuclear pore complex binding domain with NXF1, which is absent in NXF3 (Supplementary Figure 2). To characterize the interaction of NXT2 with testis-specific NXF2 and NXF3 in detail, we queried which of these domains mediates the binding to NXT2. Accordingly, we generated FLAG-tagged expression constructs of NXF2 and NXF3, lacking the NTF2-like domains or the RRM domains (Figure 2a), overexpressed them in HEK293T cells (Supplementary Figure 3a), and performed Co-IP analyses. Binding to NXT2 was abolished when the NTF2-like domain was deleted in NXF2 (Figure 2b). However, NXF2 lacking the RRM domain was still able to bind to NXT2 (Figure 2c). In line, deletion of the NTF2-like domain in NXF3 (Figure 2d; Supplementary Figure 3b) inhibited binding to NXT2 (Figure 2e), whereas deletion of the RRM domain (Figure 2d; Supplementary Figure 3b) did not affect binding capacity (Figure 2f). Besides, a C-terminally truncated NXT2, lacking the last six amino acids but containing the NTF2-like domain (Supplementary Figure 4a) was still able to bind to both NXF2 (Supplementary Figure 4b) and NXF3 (Supplementary Figure 4c). These data show that the binding of NXT2 and NXF2/3 depends on the NTF2-like domain of the respective interaction partners, while the RRM domain is dispensable *in vitro*.

**Figure 2:**
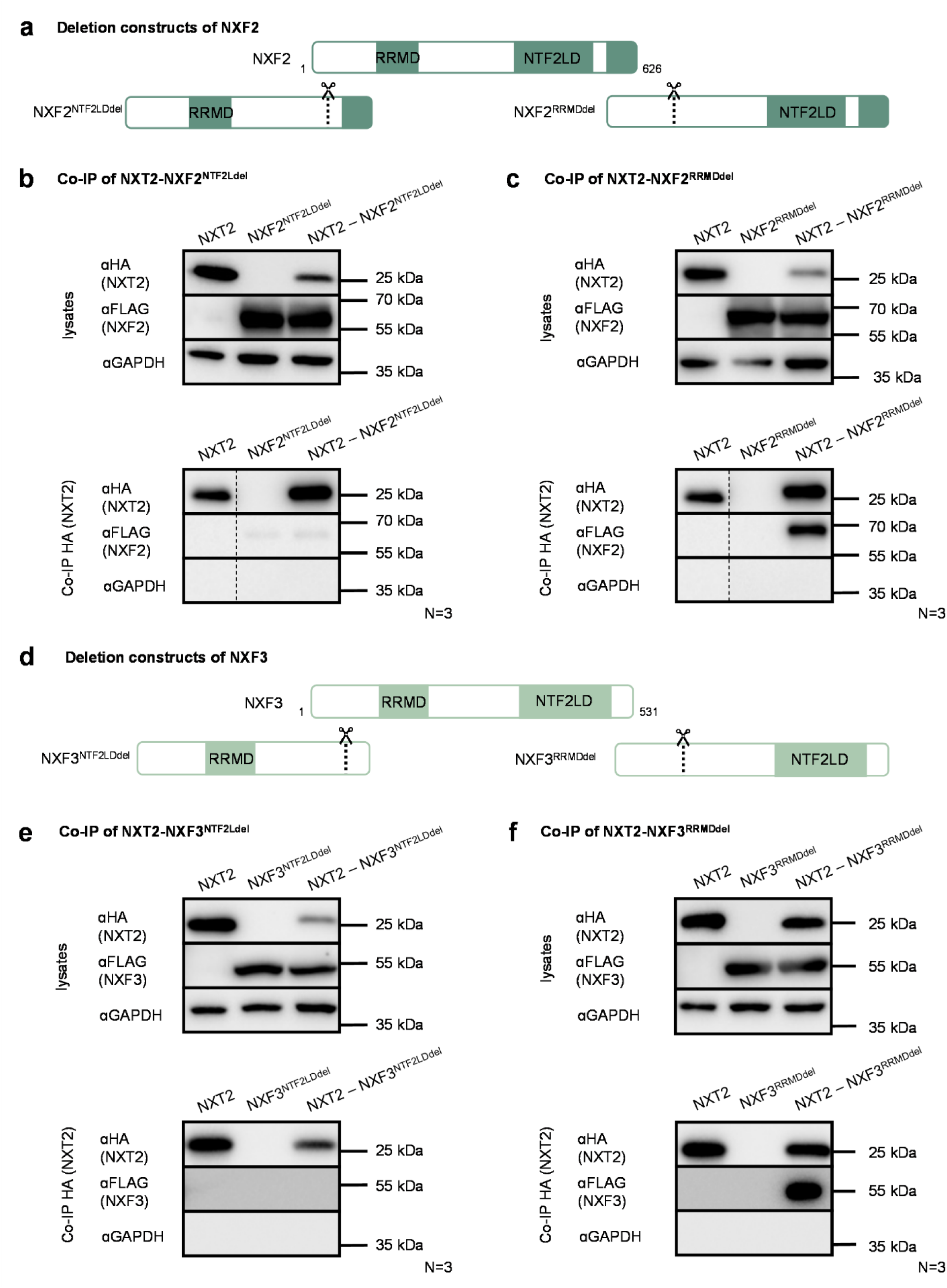
NXT2 binds to NXF2 and NXF3 by their NTF2-like domains. a. To scrutinize the protein domain mediating binding to NXT2, deletion constructs of NXF2-FLAG lacking the NTF2-like domain (NXF2^NTF2LDdel^) or the RNA recognition motif domain (NXF2^RRMDdel^) were generated. b. Western blot analyses demonstrating specific expression of NXT2 and NXF2^NTF2LDdel^ in protein lysates isolated from transfected HEK293T cells (top). In Co-IP analysis, NXF2^NTF2LDdel^ is not able to bind to NXT2 (bottom). c. NXF2^RRMDdel^ still binds to NXT2 (cell lysates derived from transfected HEK293T cells at the top, Co-IP at the bottom). d. Schematic depiction of deletion constructs of NXF3-FLAG lacking the NTF2-like domain (NXF3^NTF2LDdel^) or the RRM domain (NXF2^RRMDdel^). e. In Co-IP analysis, NXT2-NXF3 heterodimerization is dependent on the NTF2-like domain in NXF3. f. In Co-IP analysis of NXT2 and NXF2^RRMDdel^, binding of both proteins is still possible, indicating that the RRM domain is dispensable for dimerization. Representative Western blots of at least three replicates are shown. Uncropped images of all Western blots depicted are provided in the Source data file.

### Deleterious variants in NXT2 are associated with azoospermia and loss of germ cells

Since NXT2 has a testis-enriched expression profile and may have evolved testis-specific functions, we next addressed the question of whether impaired NXT2 function in the human testis impairs sperm production and causes male infertility. A first indication comes from population genetic data: NXT2’s LoF observed/expected (o/e) fraction is zero with a LoF o/e upper bound fraction (LOEUF) of 0.51 (gnomAD, v2.1.1, not yet available for v.4.1, Supplementary Table 3). Such low o/e fractions are rare and specify intolerance to LoF variants, supporting an important biological relevance of the encoded protein and selective pressure on genetic variants affecting protein function. Indeed, *TEX11* is one such X-chromosomal gene with an o/e fraction of zero and one of the best established male infertility genes^13,14^. We, therefore, screened for potentially deleterious genetic variants in *NXT2* in exome/genome sequencing data of more than 2,700 well-characterized infertile men from the Male Reproductive Genomics (MERGE) cohort. Focusing on rare (minor allele frequency [MAF] ≤0.001, gnomAD v2.1.1) LoF or high impact missense variants (with CADD score ≥10) and copy number variations in *NXT2,* we identified two infertile men. Subject M3065 showed a hemizygous single nucleotide duplication (c.354dup), resulting in a premature stop codon p.(Asp119*) (Figure 3a, Supplementary Table 3), and M2004 the single nucleotide substitution c.268G>T resulting in the substitution of an alanine to serine at position 90 in the NTF2-like domain of the NXT2 protein sequence (p.(Ala90Ser)) (Figure 3a, Supplementary Table 3). In addition, a further LoF variant in *NXT2* was identified in exome data of a second cohort of 667 infertile men from Nijmegen/Newcastle^15^. The respective subject RU00584 was positive for a large deletion encompassing the entire *NXT2* gene (Figure 3a, Supplementary Table 3). This variant had been listed in the supplemental data of a recent publication without presenting clinical or functional data^15^. Andrological evaluation in all three men concordantly described azoospermia and elevated FSH levels (Supplementary Table 4), indicating impaired spermatogenesis.

**Figure 3:**
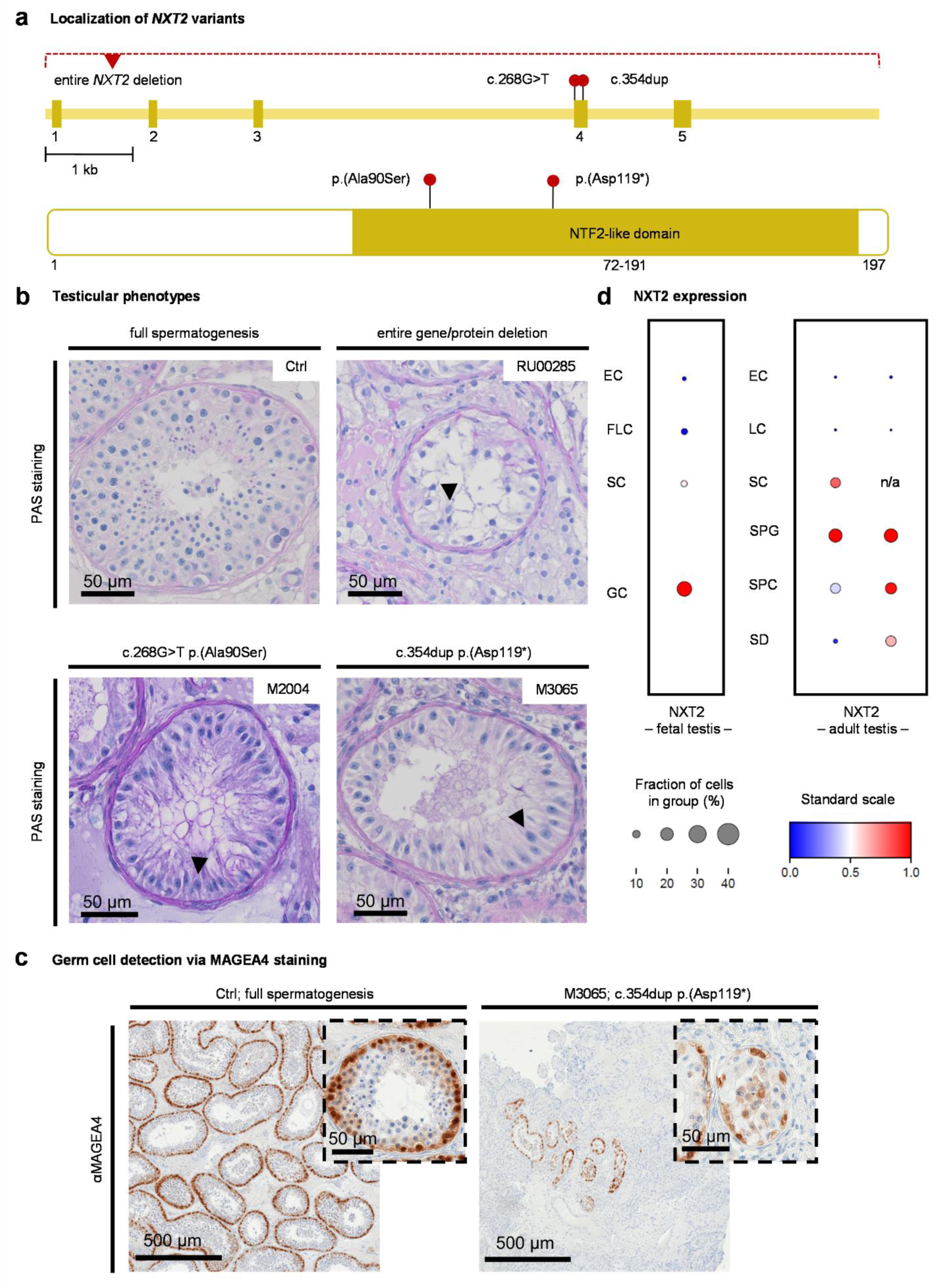
Loss-of-function variants in *NXT2* are associated with male infertility due to disturbed germ cell development. a. Schematic depiction of the *NXT2* exon-intron-structure and the NXT2 primary protein structure with the position of NTF2-like domain highlighted. Red dots indicate the localization of the identified *NXT2* variants at the gene and protein level. The missense variant p.(Ala90Ser) identified in M2004 and the LoF variant p.(Asp119*) identified in M3065 are both located within the NTF2-like domain. The deletion identified in RU00584 encompasses the entire *NXT2* gene. b. Overview of the testicular phenotype of all three men with variants in *NXT2* (PAS staining of testicular sections) compared to a control with full spermatogenesis. All subjects consistently had a Sertoli cell-only phenotype, i.e. a complete lack of germ cells, in the vast majority of seminiferous tubules. Sertoli cells are exemplary marked by arrows. c. Immunohistochemical staining for germ cell marker protein MAGEA4 reveals only sparse seminiferous tubules with few spermatogonia and spermatocytes only in M3065. No haploid germ cells were identified. d. In fetal male germ cells (left), NXT2 is highly expressed in germ cells according to scRNA-seq data. In the adult testis (right), NXT2 is mainly expressed in spermatogonia but also in Sertoli cells and in all germ cell stages up to spermatids in two different data sets (EC: endothelial cells; FLC: fetal Leydig cells; LC: Leydig cells; SC: Sertoli cells; GC: germ cells; SPG: spermatogonia; SPC: spermatocytes; RS: round spermatids).

To investigate the impact of disturbed NXT2 function on testicular germ cell development, we detailedly analyzed the subjects’ clinical and testicular phenotypes, as well as the cellular expression pattern of *NXT2* in the human testis. Periodic acid-Schiff (PAS) staining of testicular sections from all three cases, derived from testicular biopsies, revealed a common phenotype characterized by seminiferous tubules predominantly lacking germ cells (Sertoli cell-only, SCO, HP:0034299, Figure 3b). To reinforce these histological findings, we performed immunohistochemical staining for the germ cell/spermatogonial stem cell marker MAGEA4. Interestingly, only M3065 had a few focal seminiferous tubules with spermatogonia and spermatocytes (Figure 3c), but no sperm could be retrieved by testicular sperm extraction (TESE). In contrast, MAGEA4 staining was negative in M2004 and RU00584, although a few morphologically abnormal sperm were detected in a TESE-derived testicular cell suspension in RU00584. Due to their abnormal appearance, the sperm were deemed unsuitable for ICSI (Supplementary Table 4).

Published scRNA-seq datasets^16,17^ of the human fetal male germ cells and adult testis depict a strong expression of *NXT2* not only in adult but also in fetal germ cells (Figure 3d), supporting a function of NXT2 already at fetal/embryonic stages of germ cell development. In addition to germ cells, in adult testicular tissue, *NXT2* was also expressed in somatic Sertoli cells^17–19^, indicating not only a developmental, but also a cell type-specific function of NXT2 in testicular tissue (Figure 3d).

To address the genetic and functional impact of the identified variants in *NXT2*, we performed co-segregation analyses within the families and in-depth functional characterization of the variants at the protein level. M3065 shared the *NXT2* LoF variant c.354dup p.(Asp119*) with two further infertile male siblings with azoospermia (M3065B1/B3). The variant was inherited from the heterozygous female parent and was absent in fertile male relatives, thus co-segregating with azoospermia/infertility (Figure 4a). Other genetic causes were excluded by filtering for shared rare (MAF ≤0.01), coding variants in the exome data of all affected family members that were absent in unaffected male family members. In this shared variant analysis, additional rare variants, predicted to result in protein sequence alterations and additionally co-segregating with the phenotype, were identified exclusively in genes that were expressed in other tissues or ubiquitously expressed (Supplementary Table 5).

**Figure 4:**
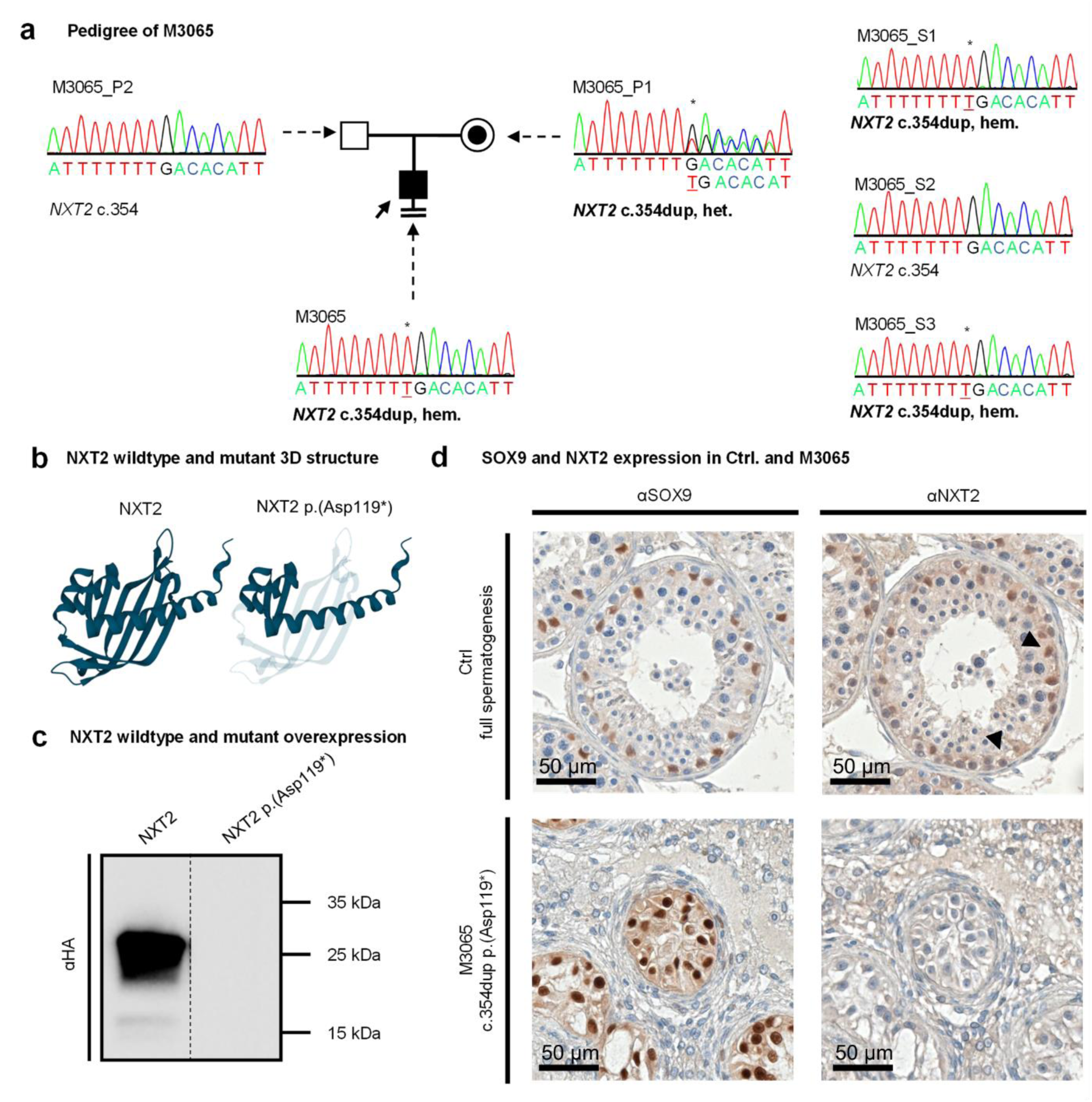
The *NXT2* variant c.354dup p.(Asp119*) co-segregates with azoospermia and leads to NXT2 deficiency. a. Index subject M3065 carries a duplication of a thymine at position 354 of the *NXT2* open reading frame resulting in the direct inclusion of a premature stop codon. The variant is inherited from the female parent (P1, heterozygous carrier) and is also present in two male siblings (S1, S3) with azoospermia. The variant is absent in the only fertile male sibling (S2). b. AlphaFold2 prediction of the short isoform (NM_001242617.2) of wildtype NXT2 (left). Inclusion of a premature stop codon results in the loss of 78 amino acids at the protein C-terminus (transparent, right). c. In Western blot analysis of protein lysates derived from overexpression of mutant NXT2-HA, no protein was detectable. d. Sertoli cells show positive SOX9 staining in control’s and M3065’s testicular tissue. In contrast, NXT2-specific staining, which is mainly visible in Sertoli cells and spermatogonia in the control tissue (as exemplary indicated by arrows; top), is absent in the subject (bottom).

The c.354dup variant introduces a premature stop codon at position 119 of the *NXT2* open reading frame, likely resulting in the degradation of the mutant mRNA by nonsense mediated decay (NMD). If, however, the transcript escapes NMD, the truncated protein would lack 78 C-terminal amino acids and thus most of the NTF2-like domain (Figure 4b). Overexpression of HA-tagged NXT2 c.354dup in HEK293T cells resulted in the complete absence of the truncated protein according to Western blot analysis (Figure 4c), arguing against the expression of a stable truncated protein. To corroborate the suspected absence of NXT2 in the subject’s testicular tissue, we performed immunohistochemical staining for NXT2 using an antibody that is directed against an NXT2 protein region that would be present in the truncated protein (Supplementary Table 1). Indeed, no NXT2-specific staining could be detected in the subject’s Sertoli cells, which were clearly assignable by positive staining for the Sertoli cell marker SOX9. In contrast, a tissue with complete spermatogenesis displayed positive staining for NXT2 in spermatogonia and Sertoli cells (Figure 4d).

RU00584 was previously described as carrying a deletion of *NXT2* identified by exome sequencing^15^, but this variant had not been not prioritized for follow-up at that time. Analysis of subsequently produced genome sequencing data indicated that the subject presents a 42 kb large deletion on the X chromosome encompassing the entire *NXT2* gene. Interestingly, the deletion occurred *de novo*, as it was not detected in the subject’s parents, providing further genetic evidence that the variant is pathogenic (Figure 5a). Fittingly, no NXT2-specific staining could be detected in Sertoli cells, identified by SOX9-positive staining, confirming the expected absence of NXT2 *in vivo* (Figure 5b).

**Figure 5:**
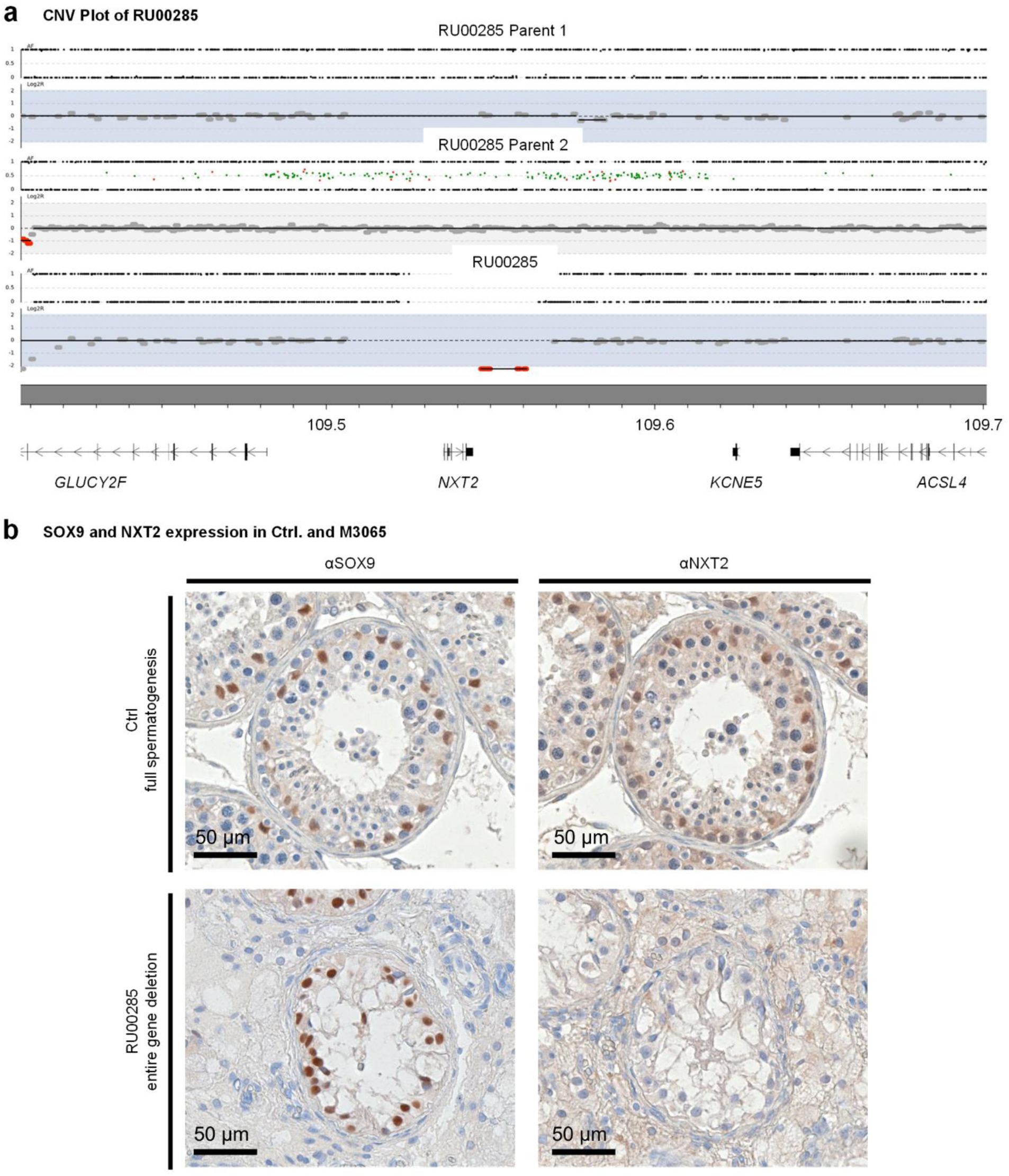
Deletion of the entire *NXT2* gene in RU00584 occurred *de novo* and results in the absence of NXT2 *in vivo*. a. Robot (CNV) plot of genome sequencing data illustrating the presence of a 42 kb deletion in RU00584 encompassing the entire *NXT2* gene and surrounding genomic regions. The deletion is absent in both parents, demonstrating *de novo* occurrence in the subject. b. *In vivo*, immunohistochemical staining for the Sertoli cell marker protein SOX9 confirms the presence of Sertoli cells in the control (top) and in RU00584 (bottom). In an immunohistochemical staining for NXT2, in contrast to the control, no NXT2-specific staining was observed in Sertoli cells in RU00584.

The third *NXT2* variant identified in M2004, c.268G>T, affects the 5’-end nucleotide of *NXT2* exon four (Supplementary Figure 5a). As this variant might affect splicing, we performed a minigene assay to analyze the effect on the recognition of the respective splice site *in vitro*. In contrast to the wildtype, the mutant *NXT2* minigene revealed two distinct splice products. In addition to the clearly visible normally spliced transcript, a faint signal was detected corresponding to an aberrantly spliced transcript lacking exon four. Skipping of this exon would result in a shift of the open reading frame and subsequent insertion of a premature stop codon (c.268G>T r.268_412del p.(Ala90Serfs*13)) (Supplementary Figure 5b). The predicted amino acid substitution resulting from c.268G>T affects the alanine residue at position 90, which is highly conserved in orthologous proteins (Supplementary Figure 5c) and positioned in a central protein domain, as indicated by the AlphaFold2 model (Supplementary Figure 5d). Because the residue is located in the NTF2-like domain, which is crucial for binding to NXF2 and NXF3, we next analyzed whether the amino acid substitution affects protein stability or binding to NXF2 or NXF3. When overexpressing mutant NXT2 in HEK293T cells, the protein expression level was comparable to the wildtype (Supplementary Figure 5e) and, in Co-IP, binding of mutant NXT2 to NXF2 (Supplementary Figure 5f) and NXF3 (Supplementary Figure 5g) was unaffected. In addition, NXT2 staining was present in SOX9-positive Sertoli cells in the subject’s testis (Supplementary Figure 5h). Accordingly, no clear functional link could be established between this missense variant and the proband’s Sertoli cell-only phenotype.

### An NXF3 variant leads to oligoasthenoteratozoospermia

To unravel whether the testis-expressed binding partners of NXT2, the nuclear export factor proteins NXF1, NXF2, and NXF3, are also important for male fertility, we screened the exome/genome data of the MERGE and Nijmegen/Newcastle cohorts for rare, potentially deleterious variants in *NXF1*, *NXF2*, and *NXF3* using the same filtering criteria as outlined above for *NXT2*.

A potentially deleterious variant was detected only in *NXF3.* One man with infertility (M2799) identified in the MERGE cohort was positive for the hemizygous stop-gain variant c.826G>T p.(Gly276*). With a LoF o/e fraction of 0.33 and a LOEUF of 0.6 (gnomAD, v2.1.1) (Supplementary Table 3) also *NXF3* indicates a reduced tolerance to LoF variants in the general population. The variant is located in exon nine of *NXF3* (Figure 6a) and would lead to a protein lacking the NTF2-like domain (Figure 6b) if the mutant transcript escaped NMD. The variant was inherited from the heterozygous female parent and was not identified in two fertile male relatives (Figure 6c). No further high-impact variants in genes expressed in the testis were present in the proband’s exome. In Western blot analyses of lysates overexpressing the mutant protein, a ∼30 kDa smaller protein (Figure 6d) compared to the wildtype protein was detected. This truncated protein was unable to bind to NXT2, as demonstrated by Co-IP (Figure 6e). Thus, even if the mutant transcript would escape NMD, the stop-gain variant would result in an abolished interaction with NXT2.

**Figure 6:**
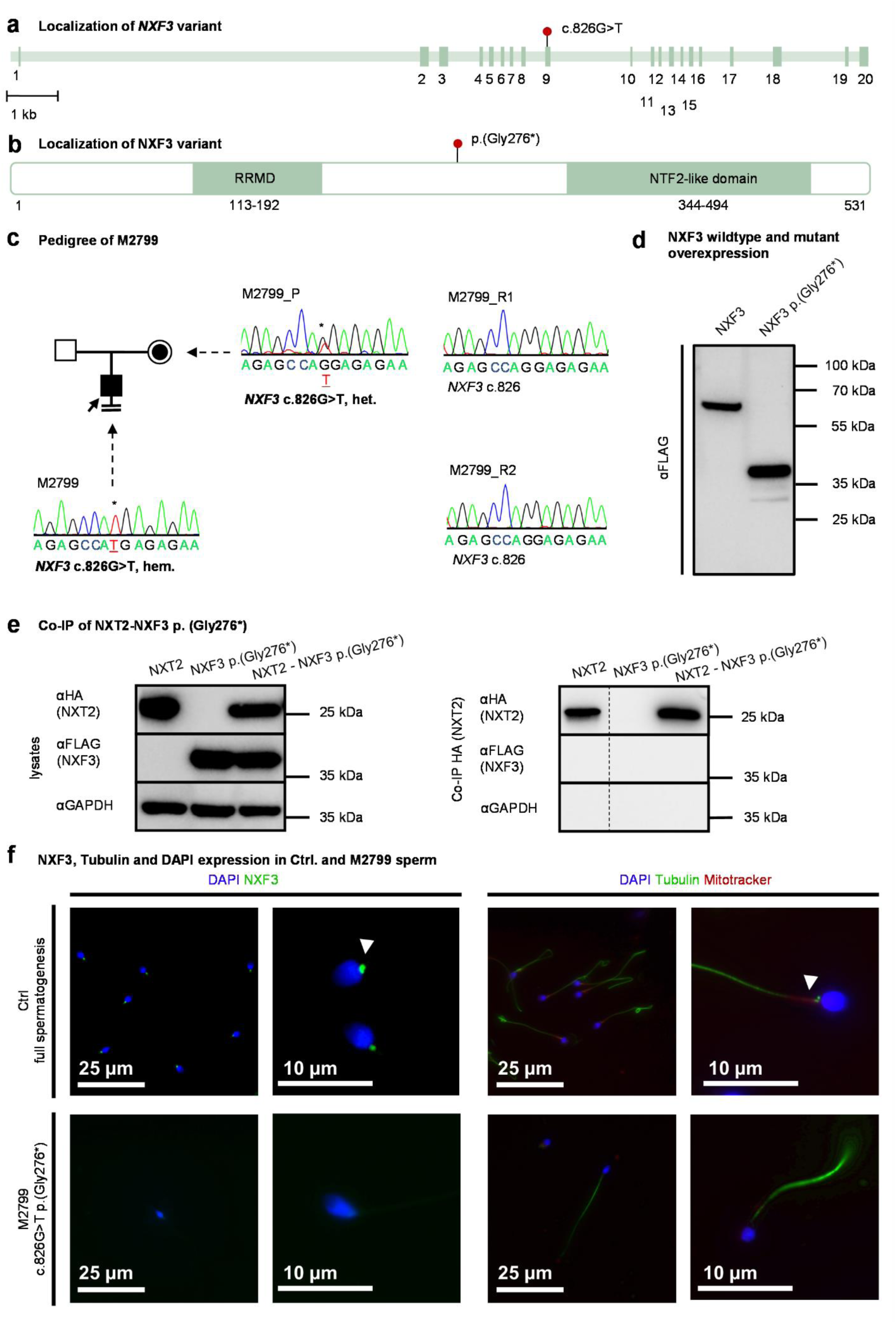
A LoF variant in *NXF3* is associated with quantitatively and qualitatively severely impaired sperm production. a. Schematic depiction of the exon-intron-structure of *NXF3* with the variant c.826G>T in exon nine indicated by a red dot. b. At the protein level, the variant leads to the inclusion of a premature stop codon p.(Gly276*), resulting in a truncated protein lacking the NTF2-like domain if the mutant transcript is not degraded by NMD. c. Pedigree of M2799 demonstrating hemizygous presence of c.826G>T in the index case. The variant is inherited from the heterozygous female parent (P) and two fertile male relatives (R1, R2) are negative for the variant. d. *In vitro*, the variant leads to the expression of a C-terminally truncated NXF3-FLAG protein (expected size: ∼32 kDa). e. Western blot analysis of lysates derived from overexpressed mutant NXF3 in HEK293T cells (left). Co-IP of HA-tagged NXT2 and mutant FLAG-tagged NXF3 demonstrates abolished binding of the truncated NXF3 protein to NXT2 (right). f. Immunofluorescence staining for NXF3 and sperm tail and midpiece marker in sperm from healthy donors and M2799. In control sperm (top left), NXF3 staining localized to the neck, very closely attached to the sperm head. In the subject’s sperm (bottom left), NXF3 staining was absent, indicating absence of the protein. Sperm were characterized by staining for specific marker proteins. The head (DAPI) is stained blue, the flagellum (α/β-tubulin) is stained green and the midpiece (Mitotracker, exemplary indicated by a white arrow) is stained red (right). M2799 sperm did not show specific staining for the midpiece marker Mitotracker (right bottom).

In contrast to the subjects with *NXT2* variants, who were concordantly azoospermic, M2799 had extreme oligozoospermia (<2 million total sperm count) and mostly immotile sperm in his ejaculate (≥85 %) in two independent semen samples. Sperm morphology was not assessed during routine semen analysis. The striking difference in the phenotypes observed in men with *NXT2* variants compared to the phenotype of M2799 with the *NXF3* variant is in line with the different expression profiles of the two genes. In contrast to *NXT2*, *NXF3* is not expressed in male fetal germ cells, according to published scRNA-seq datasets (Supplementary Figure 6a). Notably, within the adult testis, *NXF3* mRNA is present in Sertoli cells and spermatids (Supplementary Figure 6b), indicating a function of the NXT2-NXF3 heterodimer at later stages of spermatogenesis.

To analyze the localization of NXF3 in sperm, we performed immunofluorescence staining using an NXF3-specific antibody (Supplementary Figure 1c/d; Supplementary Table 1). In sperm from a control donor, NXF3 localized to the midpiece, more specifically to the centriole/connecting piece (Figure 6f). In contrast, no NXF3-specific staining was detected in M2799’s sperm, indicating the absence of protein expression *in vivo*. To further screen for morphological sperm abnormalities that could potentially explain the observed sperm immotility, we stained the sperm with a Tubulin-specific antibody, which specifically highlights the flagellum, and with Mitotracker, staining mitochondria in the midpiece (Figure 6f). Interestingly, in M2799’s sperm, Mitrotracker-specific staining was not detectable, suggesting a midpiece defect/malformation or mitochondrial dysfunction, leading to the observed impairment of sperm motility. In summary, the man with the *NXF3* LoF variant had quantitatively and qualitatively severely impaired sperm production.

## Discussion

In this study, we demonstrate that testis-enriched NXT2 is indispensable for normal spermatogenesis in humans by identifying two *NXT2* variants that abolish protein expression in infertile men with azoospermia. Both subjects share a testicular phenotype of predominant Sertoli cell-only, *i.e.*, absence of germ cells, which we link to the absence of NXT2 in the testicular tissue. One of the variants co-segregates with the infertility in the family, while for the second variant a *de novo* occurrence was demonstrated, strengthening the genetic evidence for the association with the observed phenotype^15^. Indeed, NXT2 had not yet been associated with any disease and only three heterozygous LoF variants in *NXT2,* identified in females, are present in the newest gnomAD population database of >800,000 individuals (v4.1.0), indicating intolerance to LoF variants, selective pressure, and/or incompatibility with heritability. From a clinical perspective, *NXT2* is a strong candidate gene for male infertility due to azoospermia but the identification of additional patients harboring variants in *NXT2* is necessary to firmly establish a clinically valid gene-disease relationship.

Since germ cells are largely absent in affected men’s testes, we propose that NXT2 has a critical function either in somatic Sertoli cells, being crucial for orchestrating spermatogenesis by maintaining the spermatogonial stem cell niche and providing essential growth factors^20^, and/or in first steps of germ cell development, as evidenced by the enriched expression of *NXT2* in fetal male germ cells.

In contrast to NXT1, NXT2 has undergone adaptive selection during evolution^11^ and displays a testis-enriched expression only in primates, whereas the protein is ubiquitously expressed in rodents. These different evolutionary fates already point towards a tissue-specific novel function of NXT2 restricted to the primate testis. This is supported by our finding that the absence of NXT2 in humans is associated with impaired spermatogenesis, whereas in mice *Nxt2* has been demonstrated to be dispensable for fertility^21^.

Our data also support an involvement of NXT2 in the export of bulk mRNA through the nuclear pore, as we demonstrate not only an interaction with NXF1, part of the well-characterized NXT1-NXF1 export factor complex^5^, but also with the nucleoporins NUP93 and NUP214. NUP214 contains phenylalanine-glycine (FG)-repeats that are essential for binding to NXF proteins^22^ and both NUPs are involved in the transport of RNPs through the nuclear pore^23^. In addition to the evolutionarily acquired adaptations of NXT2, both the absence of NXT1 from the interactome and the severe phenotype observed in men with *NXT2* variants suggest that, in humans, the cellular function of NXT2 cannot be compensated by NXT1, pointing towards testis-specific NXT2-NXF dependent processes.

In addition to the ubiquitously expressed *NXF1*, humans encode four additional NXF genes (*NXF2*-*NXF5*), all of which are located on the X chromosome and display mainly tissue-specific expression patterns^7^. NXF2 shares an overall similar protein structure with NXF1, reveals a testis-specific expression, and associates with mRNA *in vitro*^7^. The mouse ortholog of NXF2 localizes to the nucleus or the nuclear periphery of germ cells, supporting a function in RNA export through the nuclear pore complex^24^. *Nxf2* knockout mice revealed an age-dependent depletion of spermatogonial stem cells and male infertility^25^, and a crucial function in regulating germ cell development was proposed. However, as no men harboring potentially deleterious variants in *NXF2* have been identified yet, it remains to be seen whether loss of NXF2 will also results in human male infertility.

Mammalian NXF3 lost its nuclear pore complex binding ability^7^ but gained an additional binding site for XPO1 (also called CRM1), an exportin that facilitates translocation through the nuclear pore complex^26^. Based on the tissue-specific expression of *NXF3* in the testis, it has been speculated that the NXF3-XPO1 complex mediates the transport of specific RNAs^26^. We present genetic and functional data indicating that human NXT2 binds to NXF3 in the testis and that abrogation of NXF3 function leads to impaired sperm development at later stages of spermatogenesis. In contrast to *NXT2*, *NXF3* is not expressed in fetal male germ cells but at later stages of spermatogenesis^19^, suggesting that NXF3 is dispensable for fetal functions of the NXT2-mediated RNA transport but is likely crucial during adult spermatogenesis. Fittingly, we observed that NXF3 localizes to the sperm midpiece in sperm from a control individual, while being absent in the subject hemizygous for the *NXF3* variant. Of note, and similar to *NXT2*, knockout of the NXF3 ortholog in mice does not impact spermatogenesis^27^, again pointing towards different evolutionary fates of the human and mouse genes. The cargos of NXF3-mediated transport in mammals have not been elucidated yet and XPO1 was not identified as part of the NXT2 interactome in this study. Hence, it remains to be clarified whether the infertility in the *NXF3* LoF subject is a consequence of impaired mRNA export at later stages of spermatogenesis or a consequence of other NXF3 related functions.

Examples of other RNA nuclear export-independent functions of NXF proteins have already been described for mammalian NXF2, which is involved in cytoplasmic mRNA dynamics by interacting with motor proteins such as KIF17^28^. In addition, expression of more than one NXF protein is also known from other eukaryotic lineages, including *Caenorhabditis elegans* and *Drosophila melanogaster,* and interestingly this diversification occurred independently assuming that these NXF variants might also have evolved novel molecular functions, not directly related to mRNA export^29^ as we suggest for human NXF2/3. Indeed, in *Drosophila melanogaster*, one of the NXF proteins (*Drosophila* Nxf2) triggers co-transcriptional repression of transposons in germ cells in a Piwi-dependent manner^30^. Further, the *Drosophila* NXF3 was shown to export piRNA precursors and guide them to the nuage, a germ cell specific granule, where they are processed into mature piRNAs, which are important for protecting genome integrity by silencing transposable elements^31–33^.

Besides nuclear export factors, we have also identified SPATA5 and SPATA5L1, which initiate the cytoplasmic pre-60S maturation cascade within ribosome biogenesis and trigger the release of shuttling factors^34,35^, as part of the NXT2 interactome. Since SPATA5 directly processes the pre-60S particle after transport through the nuclear pore complex and also interacts with nucleoporins^36^, the interaction with NXT2 could indicate an involvement of NXT2 in the export of ribosomal subunits. Of note, a testis-specific ribosome with a specialized nascent peptide exit tunnel facilitating translation during sperm formation has recently been described, and deletion of this ribosome has been associated with defective sperm formation and subfertility in mice^37^.

In summary, we introduce *NXT2* as a strong candidate gene for male infertility and demonstrate that the encoded protein is the key player in adult human testis bulk RNA nucleocytoplasmic transport by interacting with the RNA export factor NXF1 and the nuclear pore complex. NXT2 also interacts with the human testis-specific nuclear export factor proteins NXF2 and NXF3 and ribosome biogenesis factors giving rise to cellular functions independent of nucleocytoplasmic mRNA transport and/or cargo specificity in the human testis. It remains to be elucidated which specific cargos are transported in NXT2 mediated export.

## Methods

### Ethical approval

All persons included in the study gave written informed consent for the analysis of their donated material and the evaluation of their clinical data compliant with local requirements. The use of testicular tissue for pulldown analysis and the MERGE study protocol were approved by the Münster Ethics Committees/Institutional Review Boards (Ref. No. Münster: 2012-555-f-S and 2010-578-f-S). Semen samples were provided by normozoospermic donors with prior written consent according to the protocols approved by the Ethics Committee of the Ärztekammer Westfalen-Lippe and the Medical Faculty Münster (4INie, 2021-402-f-S). The study protocol of the Radboudumc outpatient clinic and the Newcastle upon Tyne Hospitals NHS Foundation Trust (Newcastle, UK) was approved by the respective Ethics Committees/Institutional Review Boards (Nijmegen: NL50495.091.14 version 5.0, Newcastle: REC ref. 18/NE/0089). All procedures were in accordance with the Helsinki Declaration of 1975. Subjects’ identifiers are not known to anyone outside of the research groups.

### Lysis of human testicular tissue

Adult human testicular tissue derived from surgery of three subjects with obstructive azoospermia (OA) (pooled) and one transgender person, who still had full spermatogenesis in testicular biopsy despite of the hormonal treatment, was stored at −80 °C prior homogenization with the TissueLyser LT (QIAGEN, Hilden, Germany) for 6 min with a 5 mm Stainless Steel Bead (QIAGEN, Hilden, Germany) in Pierce IP lysisbuffer (#87787 [Thermo Scientific™, Waltham, USA]; 50 mM Tris-HCl, 1 % Triton X-100, 5 mM EDTA, 150 mM NaCl, 0.2 mM PMSF, 1 mM DTT, 1x protease inhibitor cocktail). The samples were incubated on ice for 15 min and centrifuged at 4 °C and 14000 rcf for 10 min. The supernatant was immediately used for pulldown experiments.

### Pulldown of native proteins from testicular lysates and preparation of samples for mass spectrometry

Dynabeads Protein-G (Thermo Scientific™, Waltham, USA) were incubated with 5 µg of specific NXT2 and IgG antibodies (Supplementary Table 1) for 90 min at 4 °C. To perform the pulldown, the coupled beads were incubated with testicular lysate (∼80 mg) at 4 °C overnight. Of the 120 µl sample, 20 µl were used for subsequent Western blot analysis to check for efficiency of the pulldown and 50 µl were washed with 50 mM Tris-HCl (pH 8) three times. The beads were dried and stored at −80 °C. Four independent replicates pulled down with NXT2 (3 samples form the transgender person tissue lysate, 1 sample of pooled tissue lysates from men with obstructive azoospermia, OA [HP:0011962]) and two control replicates pulled down with IgG (2 samples from transgender person tissue lysate) were sent for mass spectrometry analyses.

### LC–ESI-MS/MS Analysis

The LC–ESI-MS/MS analyses were performed in the Proteomics Facility at the University of Turku on a nanoflow HPLC system (Easy-nLC1200, Thermo Scientific™, Waltham, USA) coupled to the Q Exactive HF (Thermo Fisher Scientific, Bremen, Germany) equipped with a nano-electrospray ionization source. Peptides were first loaded on a trapping column and subsequently separated inline on a 15 cm C18 column (75 μm x 15 cm, ReproSil-Pur 3 μm 120 Å C18-AQ, Dr Maisch HPLC GmbH, Ammerbuch-Entringen, Germany). The mobile phase consisted of water with 0.1 % formic acid (solvent A) and acetonitrile/water (80:20 (v/v)) with 0.1 % formic acid (solvent B). A 70 min gradient was used to eluate peptides (60 min from 6 % to 39 min solvent B and in 2 min from 39 % to 100 % of solvent B, followed by 8 min wash stage with solvent B). MS data was acquired automatically by using Thermo Xcalibur 4.1 software (Thermo Scientific™, Waltham, USA). An data dependent acquisition method repeated cycles of one MS1 scan covering a range of 350–1750 m/z followed by HCD fragment ion scans (MS2 scans) for the 10 most intense peptide ions from the MS1 scan. Two-sided t-test on protein abundance values was used to identify significant (p<0.05) interaction partners of NXT2 compared to IgG.

### Gene Ontology Analysis

Gene ontology analysis (http://geneontology.org)^38,39^ was performed for all genes encoding proteins siginificantly enriched in the NXT2 pulldown compared to IgG (Figure 1a) (for “biological processes” and “homo sapiens”) and processed with PANTHER (https://pantherdb.org/webservices/go/overrep.jsp)^40^ (annotation dataset: “GO biological processes complete”, test type: ‘Fisher’s Exact”, Correction: “Bonferroni”, showing results with P <0.05). GO terms were then processed with Revigo62 (http://revigo.irb.hr/_)_^41^ using the P-value and a medium (0.7) list setting (obsolete GO terms were removed, species “homo sapiens”, “SimRel” semantic similarity measure). The Revigo Table was exported and −log10(P-value) of representative GO terms (classed as representation: ‘null’) were plotted with CirGO.py63^42^ for visualization of the 2-tiered hierarchy of GO-terms.

### STRING interaction analysis

To identify protein clusters within the significantly enriched proteins, a STRING functional protein association analysis was carried out^43^. There was no filtering for active interaction sources and line thickness indicates the confidence of the interaction. K-mean clustering with a given number of three clusters was executed.

### Cloning of cDNA constructs

Human adult testis RNA (BioCat, Heidelberg, Germany) was converted to cDNA using the GoScript™ Reverse Transcriptase system (Promega, Madison, USA) according to the instructions of the manufacturer. Amplification of human cDNAs encompassing the entire open reading frame of *NXT2* (NM_018698.5), *NXF2* (NM_022053.4) and *NXF3* (NM_022052.2) was performed with PrimeSTAR Max polymerase (Takara Bio, Kusatsu, Japan) and PCR products were cloned into the mammalian expression vector pcDNA3.1(+) (Genscript, Leiden, NL). Using phosphorylated primers, an HA-tag was cloned to the N-terminus of *NXT2* and a FLAG-tag was added to the C-terminus of *NXF2* and N-terminus of *NXF3*, respectively. Deletions of respective protein domains and addition of Kozac-sequences to the 5’-end of *NXF2* and *NXF3* constructs were introduced to increase expression by PCR using phosphorylated primers and blunt end ligation of PCR products. All constructs were verified by Sanger sequencing. Primer sequences can be found in Supplementary Table 6.

### Mutagenesis of constructs

Point mutations identified in infertile men were introduced using site-directed-mutagenesis according to manufacturer’s instructions (QuikChange II XL Site-Directed Mutagenesis Kit, Agilent Technologies, Santa Clara, USA). The *NXT2* LoF variant (c.354dup), the *NXT2* missense variant (c.268G>T) and the *NXF3* LoF variant (c.826G>T) were introduced in wildtype (WT) *NXT2* and WT *NXF3* cDNA clones, respectively. Successful mutagenesis was verified by Sanger sequencing (see Supplementary Table 6 for primer sequences).

### Culture and transfection of HEK293T cells

Human embryonic kidney (HEK) 293T cells were cultured in Dulbecco’s Modified Eagle Medium (DMEM, Sigma-Aldrich, Munich, Germany), supplemented with 10 % fetal calf serum (FCS) and 1 % penicillin-streptomycin. The cells were maintained in T75 cell culture flasks at 37 °C and 5 % CO_2_. Passaging was accomplished twice a week and cells were used up to passage 20. Per well 400,000 cells were seeded into 6-well plates and transfected the following day using K2 transfection reagent (Biontex Laboratories GmbH, München, Germany) with either 2 µg (single plasmid transfections) or 4 µg plasmid DNA (co-transfections). For co-transfections of two constructs, the amounts of transfected DNA were harmonized according to the length of the fragments on cDNA level. A medium change was performed 6 h after transfection and protein lysates were prepared 48 h after transfection. Transfection and subsequent analyses were performed in independent triplicates.

### Lysis of HEK293T cells

Transfected HEK293T cells were detached using ice-cold phosphate buffered saline (PBS), followed by 5 min of centrifugation at 4 °C and 2000 rcf. Lysis was performed by thorough manual pipetting. Different lysis buffers were used for standard lysates (25 mM HEPES, 100 mM NaCl, 1 mM CaCl_2_, 1 mM MgCl_2_, 1 % TritonX-100, 1x protease inhibitor cocktail) and Co-IP samples (0.025 M Tris, 0.15 M NaCl, 0.001 M EDTA, 1 % NP-40, 1 % glycerol, 1x protease inhibitor cocktail). After an incubation of 15 min on ice and centrifugation for 15 min at 4 °C and 13000 rcf supernatants were separated. Samples were either directly denatured and used for Western blot or directly taken for Co-IP experiments.

### Co-IP of overexpressed proteins

HA-coupled magnetic beads (Thermo Scientific™, Waltham, USA) were used for Co-IP experiments according to manufacturer’s instructions. The IP of HEK293T cell lysates was carried out 30 min at room-temperature in a rotator, followed by three (NXT2-NXF3) or nine (NXT2-NXF2) washes with 0.05 % TBS-T and a final wash with H_2_O. An acidic elution (elution buffer pH 2.0) was performed for 8 min prior pH neutralization. For each approach both co-expressed proteins were also separately transfected and used as positive and negative controls.

### Western blot

65 µl lysate were mixed with 25 µl 4x Laemmli (Bio-Rad, Hercules, USA) and 10 µl DTT and denatured at 95 °C for 10 min. Samples were separated on Mini-PROTEAN® TGX StainFree™ Precast gels (Bio-Rad, Hercules, USA) and transferred to a PVDF membrane using a Trans-blot Turbo Mini Transfer Pack kit (Bio-Rad, Hercules, USA) according to manufacturer’s instructions. Membranes were blocked with 5 % milk powder in 0.025 % TBS-Tween (TBS-T) solution for 30 min at room temperature (RT) prior to primary antibody incubation (Supplementary Table 1) overnight at 4 °C. After washing, a peroxidase-conjugated secondary antibody incubation (2 h, RT, Supplementary Table 1) followed. Chemiluminescence was detected with the Clarity™ Western ECL Substrate kit (Bio-Rad, Hercules, USA) and the ChemiDoc MP Imaging System (BioRad, Hercules, USA). To assess and confirm the molecular weights of analyzed proteins, a PageRuler^TM^ plus prestained protein ladder (Thermo Scientific, Waltham, USA) was used.

### Study cohorts

The Male Reproductive Genomics (MERGE) cohort included data of 2,703 men (2,629 with exome and 74 with genome sequencing) mainly recruited in the Centre of Reproductive Medicine and Andrology (CeRA) in Münster with various infertility phenotypes. Most men of this cohort had azoospermia (N = 1,622, HP:0000027) or severely reduced sperm counts: N = 487 with cryptozoospermia (sperm only identified after centrifugation of the ejaculate, HP:0030974); N = 168 with extreme oligozoospermia (total sperm count <2 million, HP:0034815); N = 85 with severe oligozoospermia (sperm count <10 million, HP:0034818). History of oncologic diseases, including testicular tumors, as well as numerical chromosomal aberrations, such as Klinefelter syndrome (karyotype 47,XXY) and Y-chromosomal AZF-deletions, led to an exclusion. Likely pathogenic monogenic causes for the infertility phenotype have already been described in about 8 % of cases^14^.

One further subject with a deletion of the entire *NXT2* gene was identified among the cohort of 677 infertile men from Nijmegen/Newcastle and the respective variant has already been mentioned in the Supplementary data in a recent publication^15^.

### Exome sequencing, variant filtering, and validation of sequence variants

Sequencing for the MERGE cohort has been described previously^44^. In brief, genomic DNA was extracted from peripheral blood leukocytes via standard methods. For exome sequencing of the MERGE cohort, the samples were prepared and enrichment was carried out according to the protocol of either Agilent’s SureSelectQXT Target Enrichment for Illumina Multiplexed Sequencing Featuring Transposase-Based Library Prep Technology (Agilent) or Twist Bioscience’s Twist Human Core Exome. To capture libraries, Agilent’s SureSelect Human All Exon Kits V4, V5 and V6 or Twist Bioscience’s Human Core Exome plus RefSeq spike-in and Exome 2.0 plus comprehensive spike-in were used. For whole genome sequencing of samples from the MERGE cohort, sequencing libraries were prepared according to Illumina’s DNA PCR-Free library kit. For multiplexed sequencing, the libraries were index tagged using appropriate pairs of index primers. Quantity and quality of the libraries were determined with the ThermoFisher Qubit, the Agilent TapeStation 2200, and Tecan Infinite 200 Pro Microplate reader, respectively. Sequencing was performed on the Illumina NextSeq 500 System, the Illumina NextSeq 550 System, or the NovaSeq 6000 System, using the NextSeq 500/550 V2 High-Output Kit (300 cycles), or the NovaSeq 6000 S1 and S2 Reagent kits v1.5 (200 cycles), respectively. After trimming of remaining adapter sequences and primers with Cutadapt v1.15^45^, reads were aligned against GRCh37.p13 using BWA Mem v0.7.17^46^. Base quality recalibration and variant calling were performed using the GATK toolkit v3.8^47^ with haplotype caller according to the best practice recommendations. For more recent samples and whole genome samples Illumina Dragen Bio-IT platform v4.2 was used for alignment and variant calling. Both pipelines use GRCh37.7.p13 as reference genome. Resulting variants were annotated with Ensembl Variant Effect Predictor^48^. Exome/genome data were screened for rare (minor allele frequency [MAF] ≤0.001 in gnomAD v2.1.1) variants in *NXT2*, *NXF1*, *NXF2*, and *NXF3* with a predicted effect on protein function, including copy number variants, loss-of-function variants (frameshift, stop-gain, start-loss, splice site) and amino acid substitutions with a CADD score ≥10.

For the Nijmegen/Newcastle cohort, blood samples from probands and saliva samples from parents were used to extract DNA using the QIAGEN® Gentra® Puregene® DNA extraction kit according to manufacturer’s instructions (QIAGEN®, Venlo, NL). Samples were prepared and enriched according to manufacturer’s protocols of either Illumina’s Nextera DNA Exome Capture kit or Twist Bioscience’s Twist Human Core Exome Kit for exome sequencing, samples submitted for whole genome sequencing were prepared instead following the manufacturer’s instructions for the llumina TruSeq DNA PCR-free® library preparation kit followed. All samples were sequenced on a NovaSeq 6000 Sequencing System (Illumina, San Diego, USA). Following best practices recommendations sequencing data was processed as described previously^15^. Briefly, sequenced reads were aligned to the Genome Reference Consortium human assembly 38 (GRCh38/hg38) through BWA-MEM. Single nucleotide variations and small indels were identified and quality-filtered using GATK’s HaplotypeCaller v4.2.6.1^49^. Copy number variation (CNV) analysis of whole genome sequencing data was performed by combining Dysgu-sv^50^ and GATK-based CNVRobot (https://github.com/AnetaMikulasova/CNVRobot) with default parameters. Afterwards, CNVs predicted to be *de novo* were inspected through IGV^51^.

All identified variants were verified by Sanger sequencing; for primer sequences, see Supplementary Table 6. If available, segregation analyses were carried out with DNA from family members. PCR products were purified and sequenced using standard protocols.

### Minigene assay

To assess the impact on splicing of the missense variant c.268G>T p.(Ala90Ser) that affects the first nucleotide of exon 4 in *NXT2*, an *in vitro* splicing assay based on a minigene construct was performed. Primers flanking exon 4 of *NXT2* (Supplementary Table 6) were used to amplify the region of interest from genomic DNA of M2004 as well as a human control sample by standard PCR using 0.4 U of Phusion™ High-Fidelity DNA Polymerase (Thermo Scientific™, Waltham, USA). PCR products were cloned into pENTR™/D-TOPO® (Thermo Scientific™, Waltham, USA) according to manufacturer’s instructions. Gateway cloning was performed using Gateway™ LR Clonase™ Enzyme Mix (Thermo Scientific™, Waltham, USA) and pDESTsplice as destination vector (pDESTsplice was a gift from Stefan Stamm (Addgene plasmid #32484))^52^. A transient transfection with X-tremeGENE™ 9 transfection reagent (Sigma-Aldrich, St. Louis, USA) of Human Embryonic Kidney cells, HEK293T293T Lenti-X (Clontech Laboratories, Inc., Mountain View, USA) was carried out with mutant and wildtype *NXT2* minigenes. 24 h after transfection, total RNA was extracted using the RNeasy Plus Mini Kit (QIAGEN, Hilden, Germany) and reverse-transcribed into cDNA with the ProtoScript II First Strand cDNA Synthesis Kit (New England Biolabs GmbH, Frankfurt am Main, Germany). Amplification of the region of interest was performed using primers annealing to the rat insulin exons 3 and 4 that are part of the minigene construct (Supplementary Table 6). PCR products were separated on a 2 % agarose gel, bands were extracted using the QIAquick Gel Extraction Kit (QIAGEN, Hilden, Germany) and sequenced (Supplementary Table 6). For better visualization, PCR products were additionally analyzed on a 4150 TapeStation System (Agilent Technologies, Santa Clara, USA).

### Period acid-Schiff staining of human testicular tissue

Subjects M2004, M3065 and RU00584 underwent testicular biopsy, as indicated by diagnosis of non-obstructive azoospermia (NOA) according to EAU guidelines^53,54^ with the aim of testicular sperm extraction. After written informed consent, testicular biopsies were taken, immediately fixed in Bouin’s solution, and subsequently embedded in paraffin wax. Periodic acid-Schiff (PAS) staining was carried out according to previously published protocols^55,56^. In brief, sections were dewaxed in solvent (ProTaqs Clear, #4003011; Quartett Immunodiagnostika and Biotechnologie, Berlin, Germany), rehydrated in a decreasing ethanol series and then incubated for 15 min in 1 % periodic acid. After washing with dH_2_O sections were incubated for 45 min with Schiff’s reagent (Roth, Karlsruhe, Germany). Histological evaluation was performed following score count analysis^55^. Images were taken using an Olympus BX41 bright-field microscope equipped with a Leica DMC4500 camera.

### Histological staining of NXT2, SOX9, and MAGEA4 in subjects’ testicular tissue

For immunohistochemistry (IHC), deparaffinization was carried out using NeoClear (Merck, Darmstadt, Germany) and rehydration was done in a graded ethanol-water series. Antigen retrieval was performed at 90 °C in citrate buffer (pH 6) for 10 min. After 30 min of pre-blocking with 25 % goat serum, the primary antibody (NXT2/SOX9/MAGEA4; Supplementary Table 1) was applied and incubated in humidity chambers at 4 °C overnight. The secondary, biotinylated α-rabbit and α-mouse (ab6012; ab5886; Supplementary Table 1) antibodies were applied and incubated for 1 h at RT. Visualization was conducted using the avidin-biotin complex method for 45 min at RT (ABC solution - horseradish peroxidase (Vector Laboratories, Burlingame, USA)) followed by 3,3′-diaminobenzidine as substrate and counterstaining with hematoxylin. As a negative control, isotype rabbit IgG was included instead of the primary antibody in corresponding concentrations and an omission control was performed only using 5 % BSA/TBS instead of the primary antibody. Images were obtained using the PreciPoint O8 scanning microscope system.

### Immunocytochemistry of subject’s sperm

Cover slides were coated with poly-L-lysine and then incubated with approx. 2,000,000 control sperm per slide after swim-up as previously described^57^. As M2799’s sperm are immotile, only wash/centrifugation steps were performed. Fixation was done using 4 % PFA. After washing with 0.1 % Triton-X in PBS, cells were blocked with 1 % BSA in 0.1 % Triton-X in PBS. Primary antibody incubation was done overnight in a humidity chamber at 4 °C with specific antibodies against NXF3 and Tubulin (Supplementary Figure 1c/d, Supplementary Table 1). After washing, DAPI (#D1306 (Thermo Scientific™, Waltham, USA)) and secondary antibodies (Supplementary Table 1) were combined for 1 h incubation at RT (protected from light). In case of additional staining with midpiece marker Mitotracker Red CMXRos (#9082 (Cell Signaling Technology, Danver, USA), 500 nM), Mitotracker Red CMXRos was incubated for 1 h at RT after the secondary antibody and DAPI were washed away with PBS-Triton-X. After mounting with Dako Fluorescence Mounting Medium (#S3023 (Dako Denmark, Glostrup, Denmark)), slides were dried at RT overnight and then stored at 4 °C prior imaging. Imaging was done using the Leica DM2500 microscope and the Leica K3M camera.

### Prediction of NXT2 3D structure from AlphaFold2

3D structures of NXT2 were predicted and edited using AlphaFold2^58,59^. The short isoform of NXT2 (NM_001242617.2) is depicted, as it is the only available version at the last date of accession (04/24/24). Of note, for all other experiments and figures, the longer isoform of NXT2 (NM_018698.5) is used as the first exon which is only present in this isoform (NM_018698.5) is testis expressed according to GTEx^60^ (obtained from the GTEx Portal on 03/21/24).

### Statistics and Reproducibility

In the mass spectrometry data analysis, statistical comparisons between IgG and NXT2 sample groups were performed by two-sided Student’s t-test. Experimental replicates were performed as indicated in the respective figure legends. All reported variants were validated by Sanger sequencing. The investigators were not blinded to allocation during experiments and outcome assessment.

## Data Availability

Sequencing data of the MERGE study is available upon request to the corresponding author. Access to this data is limited for each case and specific consent of the respective samples. Sequencing data from the Nijmegen cohort have been deposited in the European Genome-phenome Archive(EGA) under the accession code EGAS00001005417 [https://ega-archive.org/studies/EGAS00001005417] and will be made available upon reasonable request for academic use and within the limitations of the provided informed consent by the corresponding author upon acceptance. Every request will be reviewed by the Newcastle University Male Infertility Genomics Data Access Committee; the researcher will need to sign a data access agreement after approval. All other data supporting the findings of this study are available from the corresponding author upon reasonable request. AlphaFold2 structure accession code for NXT2 is AF-Q9NPJ8-F1 [https://alphafold.ebi.ac.uk/entry/Q9NPJ8]. The mass spectrometry proteomics data have been deposited the ProteomeXchange Consortium via the PRIDE56 partner repository with the dataset identifier PXD052904 [https://www.ebi.ac.uk/pride/] and will be released upon acceptance of the manuscript. Genetic variants identified in NXT2 and NXF3 were submitted to ClinVar (SCV005043065-SCV005043067; [https://www.ncbi.nlm.nih.gov/clinvar/]). Source data are provided with this paper

https://ega-archive.org/studies/EGAS00001005417

https://alphafold.ebi.ac.uk/entry/Q9NPJ8

https://www.ebi.ac.uk/pride/

## Data availability

Sequencing data of the MERGE study is available from the corresponding author upon request. Access to this data is limited for each case and specific consent of the respective samples. Sequencing data from the Nijmegen cohort have been deposited in the European Genome-phenome Archive (EGA) under the accession code EGAS00001005417 [https://ega-archive.org/studies/EGAS00001005417] and will be made available upon reasonable request for academic use and within the limitations of the provided informed consent by the corresponding author upon acceptance. Every request will be reviewed by the Newcastle University Male Infertility Genomics Data Access Committee; the researcher will need to sign a data access agreement after approval. All other data supporting the findings of this study are available from the corresponding author upon reasonable request. AlphaFold2 structure accession code for NXT2 is AF-Q9NPJ8-F1 [https://alphafold.ebi.ac.uk/entry/Q9NPJ8]. The mass spectrometry proteomics data have been deposited the ProteomeXchange Consortium via the PRIDE^61^ partner repository with the dataset identifier PXD052904 [https://www.ebi.ac.uk/pride/]. Genetic variants identified in *NXT2* and *NXF3* were submitted to ClinVar (SCV005043065-SCV005043067; [https://www.ncbi.nlm.nih.gov/clinvar/]). Source data are provided with this paper.

## Acknowledgment

This study relied on data from probands who gave their permission for genomic analyses, and the authors gratefully thank all participants and their family members. We further would like to thank Sandra Laurentino, Leonie Herrmann, and Luisa Meier for critical reading of the manuscript.

This work was carried out within the frame of the Deutsche Forschungsgemeinschaft (DFG, German Research Foundation) funded Clinical Research Unit ‘Male Germ Cells’ (CRU326, project no. 329621271, grants to F.T. and N.N.). S.A.K. was supported by the ‘CareerS’ programme of the Medical Faculty Münster.

## Author Contributions

Study conceptualization: A.-K.D., B.S., F.T. Data curation: A.-K.D., B.S. Funding acquisition: F.T. Investigation: A.-K.D., A.A., L.M., G.v.d.H., C.K., O.K., S.A.K., M.J.X., J.V., S.K., N.N., N.K., B.S., F.T. Visualization: A.-K.D. Writing of original draft: A.-K.D., B.S., F.T. All authors revised and approved the final version of the manuscript.

