## Supplementary Information for "NXT2 is the key player for nuclear RNA export in the human testis and critical for spermatogenesis"

### Table of Contents

Supplementary Table 1. Antibody information.

Supplementary Table 2: Abundance values of significantly enriched proteins detected in mass spectrometry.

Supplementary Table 3: Genetic variants identified in *NXT2* and *NXF3*.

Supplementary Table 4. Clinical data of infertile men with *NXT2*/*NXF3* variants.

Supplementary Table 5. Analysis of M3065's familial exome sequencing data does not identify other variants that co-segregate and are candidates to cause the phenotype.

Supplementary Table 6. Primer information.

Supplementary Figure 1. NXT2/NXF3 antibody validation.

Supplementary Figure 2. Protein structures of NXT1, NXT2, NXF1, NXF2 and NXF3.

Supplementary Figure 3. Western blot analysis of NXF2 and NXF3 proteins expressed from deletion constructs lacking the NTF2-like domain or RRM domain.

Supplementary Figure 4. The NXT2 protein region downstream of the NTF2-like domain is not essential for binding to NXF2 and NXF3.

Supplementary Figure 5. The NXT2 missense variant c.268G>T p.(Ala90Ser) in M2004 has an impact on splicing but no identified effect on protein level.

Supplementary Figure 6. scRNA-seq shows expression of nuclear export factor proteins in fetal male germ cells and adult testis.

**Supplementary References**

**Supplementary Table 1. Antibody information.**

| **Antibody** | **Company, catalogue #, source species, clonality, LOT** | **Dilution, application, special protocol requirements** |
|---|---|---|
| **Primary Antibodies** | | |
| α- α/β-Tubulin | Cell Signaling Technology, #2148, rabbit, polyclonal, 8 | 1:250 (ICC) |
| α-FLAG | Merck, #F3165, mouse, monoclonal, SLCJ3741 | 1:1500 (Western blot) |
| α-GAPDH | Cell Signaling Technology, #5174, rabbit, monoclonal, 8 | 1:1500 (Western blot) |
| α-HA | Roche, #11867423, rat, monoclonal, 65506600 | 1:2000 (Western blot) |
| α-IgG | Cell Signaling Technology, #2729, rabbit, polyclonal, I5006 | 5 µg (pulldown of testicular lysates) |
| α-MAGEA4 | Abcam, #ab139297, mouse, monoclonal, GR3177245-18 | 1:500 in 5 % BSA/TBS (IHC) |
| α-NXF3 | Merck, #HPA046757, rabbit, polyclonal, R43903 | 1:500 (Western blot)<br>1:250 (ICC) |
| α-NXT2 | Merck, #HPA072010, rabbit, polyclonal, R102934 | 5 µg (pulldown of testicular lysates)<br>1:1000 (Western blot)<br>1:50 in 5 % BSA/TBS (IHC) |
| α-SOX9 | Sigma Aldrich, #AB5535, rabbit, polyclonal, 3481414 | 1:1000 in 5 % BSA/TBS (IHC) |
| **Secondary Antibodies** | | |
| α-mouse | Abcam, #5886, goat, monoclonal, GR195138-16 | 1:100 in 5 % BSA/TBS (IHC) |
| α-mouse IgGκ BP-HRP | Santa Cruz Biotechnology, #sc-516102, donkey, monoclonal, F1620 | 1:1000 (Western blot) |
| α-rabbit | Abcam, #ab6012, goat, monoclonal, GR3353084-9 | 1:100 in 5 % BSA/TBS (IHC) |
| α-rabbit Alexa Fluor Plus 488 | Thermo Scientific, #A32790, donkey, monoclonal, VE306220 | 1:250 (ICC) |
| α-rabbit IgG HRP | Santa Cruz Biotechnology, #sc-2357, mouse, monoclonal, E0819 | 1:1000 (Western blot) |
| α-rat HRP | Merck, #A9037, goat, monoclonal | 1:2000 (Western blot) |

**Supplementary Table 2: Abundance values of significantly enriched proteins detected in mass spectrometry.**

| | Testis tissue of one transgender person | | | | | Pooled testis tissue of biopsies of three persons with obstructive azoospermia | |
|---|---|---|---|---|---|---|---|
| **Protein** | **Abundance IgG 1** | **Abundance IgG 2** | **Abundance NXT2 1** | **Abundance NXT2 2** | **Abundance NXT2 3** | **Abundance NXT2 4** | **T-Test** |
| NXF1 | 14230731.38 | 14016786.41 | 375889307.8 | 403762086 | 388763931.2 | 411400234.9 | 1.96331E-05 |
| NXT2 | 18103574.63 | 25231497.75 | 765580724.5 | 937477492 | 912382921 | 904036323.8 | 0.000182886 |
| NXF3 | 22481439 | 23742693.63 | 1082563670 | 1182345274 | 1165130874 | 954071612.1 | 0.000249814 |
| NUP214 | 27171387.25 | 26968132.5 | 156840915.1 | 176367369.6 | 192071882.8 | 148902529.1 | 0.000705988 |
| NXF2 | 36633030.38 | 27647722.47 | 146708530.4 | 143763023 | 122286826.6 | 108888490.3 | 0.000658747 |
| SPATA5 | 7206384.125 | 6383698.625 | 69633338.38 | 104406044.5 | 80504325.5 | 96207154.25 | 0.001873179 |
| SPATA5L1 | 3749882.688 | 5475614.438 | 31508764.69 | 50992627.25 | 46634457.06 | 54747581.31 | 0.003342401 |
| NUP98 | 11591541.44 | 15308999.5 | 24356397.69 | 28445671.75 | 32517270.75 | 38240065.5 | 0.007674736 |

**Supplementary Table 3: Genetic variants identified in *NXT2* and *NXF3*.**

| Subject ID | Gene | Variant g. | Variant c. | Variant p. | Genotype | MAF (gnomAD v2.1.1/ v4.1.0) | LoF o/e fraction (gnomAD v2.1.1) | LoF o/e upper bound fraction (LOEUF) (gnomAD v2.1.1) |
|---|---|---|---|---|---|---|---|---|
| M2004 | *NXT2* | chrX:108,784,704G>T | c.268G>T | p.(Ala90Ser) | hem. | 0.00002909 /0.0001080 | 0 | 0.51 |
| M3065 | *NXT2* | chrX:108,784,790dup | c.354dup | p.(Asp119*) | hem. | - | | |
| RU00584 | *NXT2* | chrX:108,765,361-108,807,493del | | | hem. | - | | |
| M2799 | *NXF3* | chrX:102,337,247G>T | c.826G>T | p.(Gly276*) | hem. | - | 0.33 | 0.6 |

**Supplementary Table 4: Clinical data of infertile men with *NXT2*/*NXF3* variants.**

| Individual | Genotype | Andrological phenotype | Testicular phenotype (PAS/MAGEA4), TESE outcome |
|---|---|---|---|
| M2004 | *NXT2*:<br>g.108,784,704<br>c.268G>T<br>p.(Ala90Ser) | FSH: 9.4<br>LH: 3.2<br>T: 15.4<br>TV: 20/20<br>Azoospermia | 98 % SCO, 2 % TS/<br>no germ cells detected<br>no sperm retrieved |
| RU00584 | *NXT2*:<br>g.108,765,361-<br>108,807,493del[1] | FSH: 14.9<br>LH: 4.7<br>T: 9.6<br>TV: 10/12<br>Azoospermia | SCO/no germ cells detected<br>no sperm retrieved, some spermatozoa seen during inspection of biopsy-derived cell suspension - due to their very abnormal morphology material was not cryopreserved |
| M3065 | *NXT2*:<br>g.108,784,790dup<br>c.354dup<br>p.(Asp119*) | FSH: 63.8<br>LH: 9.7<br>T: 12.7<br>TV: 3/3<br>Azoospermia | 70 % SCO, 30 % TS/<br>focal spermatogenesis in ~7 % (11/161) of seminiferous tubules<br>no sperm retrieved |
| M2799 | *NXF3*:<br>g.102,337,247<br>c.826G>T<br>p.(Gly276*) | FSH: 4.7<br>LH: 5.8<br>T: 20.5<br>TV: 17/18<br>Oligoasthenoteratozoospermia | no biopsy performed |

TESE: testicular sperm extraction, FSH: follicle stimulating hormone (IU/L), LH: luteinizing hormone (IU/L), T: testosterone (nmol/L), TV: testicular volume right/left (mL), SCO: Sertoli cell-only, TS: tubular shadows | Reference values: FSH 1-7 IU/L, LH 2-10 IU/L, T >12 nmol/L, TV >12 mL per testis.

**Supplementary Table 5. Analysis of M3065's familial exome sequencing data does not identify other variants that co-segregate and are candidates to cause the phenotype.**

| Gene | Chr. | Variant c. | Variant p. | Consequence | MAF (gnomAD) | CADD |
|---|---|---|---|---|---|---|
| ELF4 | X | c.1810C>T | p.(Arg604Cys) | missense variant | 0.26 % | 20.7 |
| NLGN3 | X | c.63C>A | p.(Ser21Arg) | missense variant | 0.01 % | 17.35 |
| RIPPLY1 | X | c.362A>G | p.(Asn121Ser) | missense variant | 0.08 % | 19.26 |
| TIMP1 | X | c.195G>T | p.(Met65Ile) | missense variant | 0.03 % | 18.83 |

**Supplementary Table 6: Primer information.**

| | |
|---|---|
| **Sanger Sequencing** | |
| *NXT2* c.268G>T | |
| Forward | 5’-CCAGTGGAGATGGAGCAAGG-3’ |
| Reverse | 5’-ATCCAGCCCTGAAACAGCAT-3’ |
| *NXT2* c.354dup | |
| Forward | 5'-CCAGTGGAGATGGAGCAAGG-3' |
| Reverse | 5'-GTTTCTCGCTGCTTTTGGTGT-3' |
| *NXF3* c.826G>T | |
| Forward | 5'-TGCAGAAGGGCGCTATCAAA-3' |
| Reverse | 5'-AGAAAAAGGGGGAGGAGGGT-3' |
| **Minigene assay** | |
| *NXT2* c.268G>T (Initial amplification) | |
| Forward | 5’-CACCACACTAAAACTGGCTTGGGG-3' |
| Reverse | 5'-GACATTTCAGGGTCCAGCAG-3' |
| Minigene exon primers (Rat *INS2* exons 3 and 4) | |
| Forward | 5'-CCTGCTCATCCTCTGGGAGC-3' |
| Reverse | 5'-AGGTCTGAAGGTCACGGGCC-3' |
| **Cloning of cDNA constructs** | |
| *NXT2* (NM_018698.5) | |
| Forward | 5’-GGTGGTAAGCTTATGAGAAAATACAGAAGCCACTGGTC-3’ |
| Reverse | 5’-GGTGGTCTCGAGTTAACTACTAGACCAATCTTGAAAACGG-3’ |
| *NXF2* (NM_022053.4) | |
| Forward | 5’-GGTGGTAAGCTTATGTGCTCTACTCTAAAGAAGTGTGGG-3’ |
| Reverse | 5’-GGTGGTCTCGAGTTAGGAGATTTGCTTGAAGGCCTCCGCG-3’ |
| *NXF3* (NM_022052.2) | |
| Forward | 5’-GGTGGTGAATTCATGTCACTGCCTTCAGGACACACTACG -3’ |
| Reverse | 5’-GGTGGTCTCGAGTTACGAAGGCAGCATTTTCTGCTGCTCC-3’ |
| **Primers for cloning of 5’ Kozac sequence (phosphorylated)** | |
| *NXF2* | |
| Forward | 5’-ATGTGCTCTACTCTAAAGAAGTG-3’ |
| Reverse | 5’-GGTGGCAAGCTTAAGTTTAAACGCTAGCCA-3’ |
| *NXF3* | |
| Forward | 5’-ATGGACTACAAAGACGATGACGA-3’ |
| Reverse | 5’-GGTGGCGAATTCCACCACACTGGACTAGTG-3’ |
| **Primers for cloning of tags and for introducing deletions (phosphorylated)** | |
| *NXT2* (HA tag) | |
| Forward | 5’-AGAAAATACAGAAGCCACTGGTC-3’ |
| Reverse | 5’-AGCGTAATCTGGAACATCGTATGGGTACATAAGCTTAAGTTTAA-ACGCTAG-3’ |
| *NXF2* (3xFLAG tag) | |
| Forward | 5’-CACGACATCGACTACAAGGACGACGACGACAAGTGATAAGTCTA-GAGGGCCCGTTTAAACC-3’ |
| Forward 2 | 5’-GACTACAAGGACCACGACGGTGACTACAAGGACCACGACATCG-ACTACAAGGACGACG-3’ |
| Reverse | 5’-GGAGATTTGCTTGAAGGCCTCC-3’ |

| | |
|---|---|
| *NXF3* (FLAG tag) | |
| Forward | 5’-TCACTGCCTTCAGGACACACTAC-3’ |
| Reverse | 5’-CTTGTCGTCATCGTCTTTGTAGTCCATGAATTCCACCACACTGG-ACTA-3’ |
| *NXT2* (AA137-142del) | |
| Forward | 5’-AAAACGGAAGCAATCACTTGCAA-3’ |
| Reverse | 5’-TAACTCGAGTCTAGAGGGCCCGT-3’ |
| *NXF2* (NTF2LD_del) | |
| Forward | 5’-AGGGATGCCAGCCCCCAAGAGAC-3’ |
| Reverse | 5’-ATGCTTTAGGGTCTCAGATCCAG-3’ |
| *NXF3* (NTF2LD_del) | |
| Forward | 5’-CGGGATACCAGCCACCAAGGGAC-3’ |
| Reverse | 5’-ATTCTTCAACATCTCAGATCCAA-3’ |
| *NXF2* (RRMD_del) | |
| Forward | 5’-TCTGTGAAGAATAAGTTGAAGCC-3’ |
| Reverse | 5’-GTTCCTTGTGTATCCATCCTGTG-3’ |
| *NXF3* (RRMD_del) | |
| Forward | 5’-TTTGTGCACAGGGAGCTGAAGTC-3’ |
| Reverse | 5’-GCTCCCTAAGGTCCCATCCGGCA-3’ |
| **Primers for site-directed-mutagenesis** | |
| *NXT2* (c.268G>T) | |
| Forward | 5’-CGGTCACTAACCAGGCTGTATCTGGAC-3’ |
| Reverse | 5’-GGTTAGTGACCGTCTTCTTTTATCCATTGTCTC-3’ |
| *NXT2* (c.354dup) | |
| Forward | 5’-CTAAATAATTTTTTTTGACACATTGCCTTCTAGTGAGTTC-3’ |
| Reverse | 5’-GTGTCAAAAAAAATTATTTAGGGCATCCAGCCCTG-3’ |
| *NXF3* (c.826G>T) | |
| Forward | 5’-AGCCATGAGAGAAGTGTGCGGACAGAAGCC-3’ |
| Reverse | 5’-ACTTCTCTCATGGCTCTATCCCTTTCCACTTGTC-3’ |

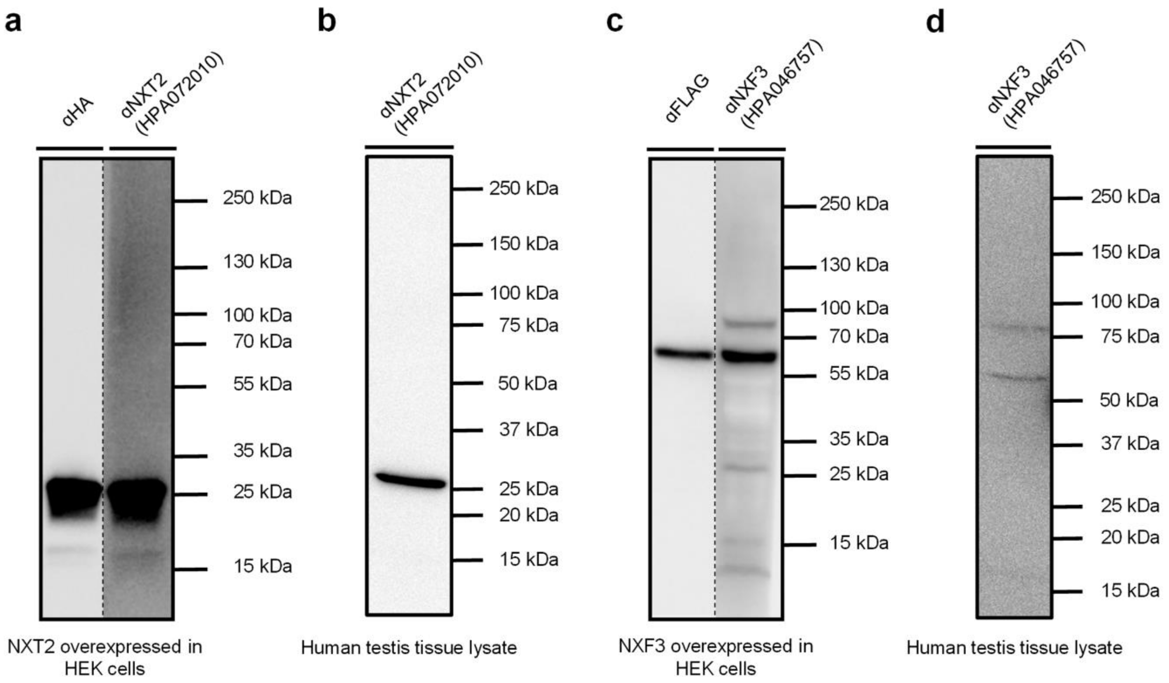


**Supplementary Figure 1. NXT2/NXF3 antibody validation.**

a. Western blot analysis of protein lysates isolated from HEK cells transfected with HA-tagged NXT2 indicating specific staining of the recombinant NXT2 protein with both HA-tag antibody and NXT2 antibody (HPA072010). Both approaches resulted in a distinct band with the expected size of ~26 kDa. b. In lysates of native human testicular tissue NXT2 was specifically detected with the NXT2 antibody (HPA072010). c. Western blot analysis of protein lysates isolated from HEK cells transfected with FLAG-tagged NXF3 indicating specific staining of the recombinant NXF3 protein with FLAG-tag antibody and NXF3 antibody (HPA072010). Both Western blots resulted in a distinct band with the expected size of ~60 kD. d. In native testicular tissue lysates, NXF3 can be detected using the NXF3 specific antibody (HPA046757). Uncropped images of all Western blots depicted are provided in the Source data file.

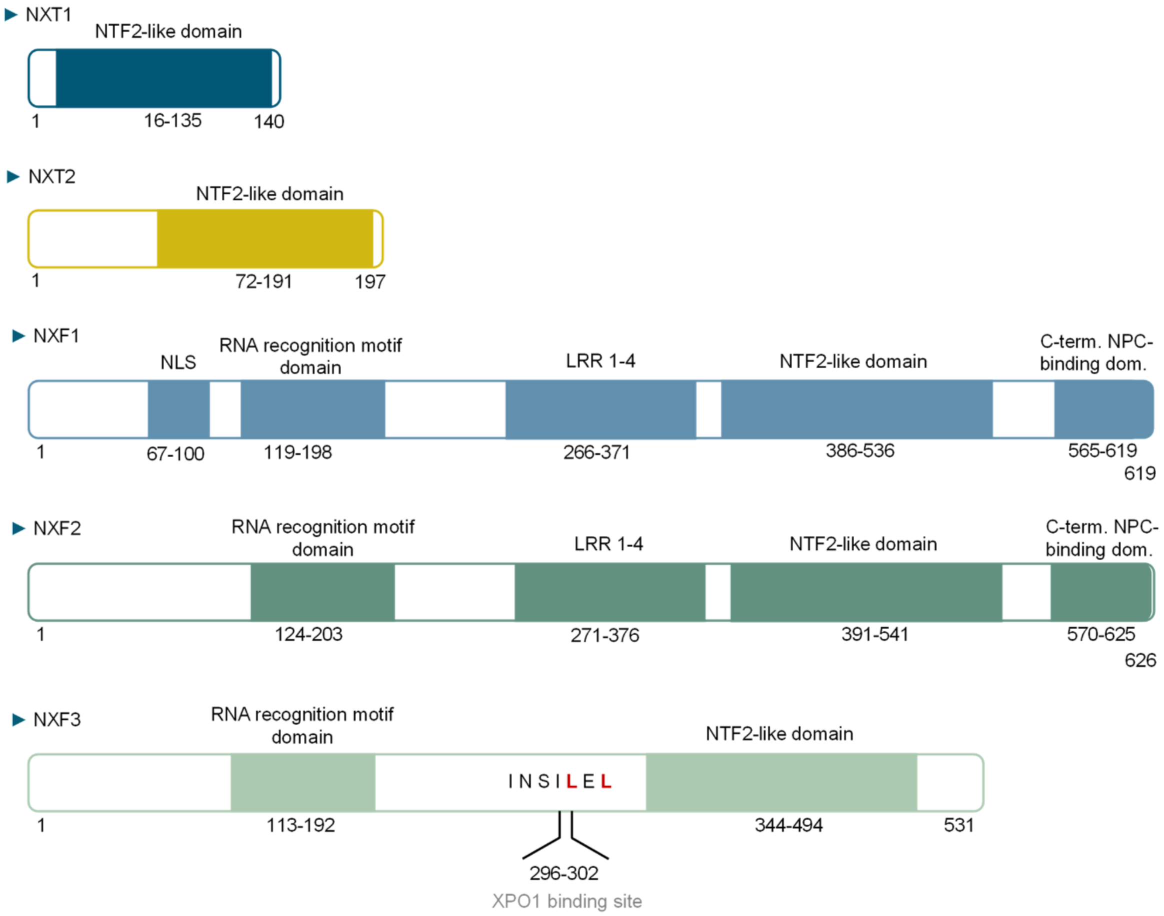


**Supplementary Figure 2. Protein structures of NXT1, NXT2, NXF1, NXF2, and NXF3.** NXT1 and NXT2 consist almost exclusively of a nuclear transport factor 2-like domain (NTF2LD). NXF1 exhibits a nuclear localization signal (NLS), an RNA recognition motif domain (RRMD), four leucine-rich repeats (LRR), an NTF2-like domain and a C-terminal nuclear pore binding domain (C-term. NPC-binding domain). NXF2 shows a similar protein structure. NXF3 is lacking the C-terminal NPC-binding domain but gained an exportin (XPO1) binding site.

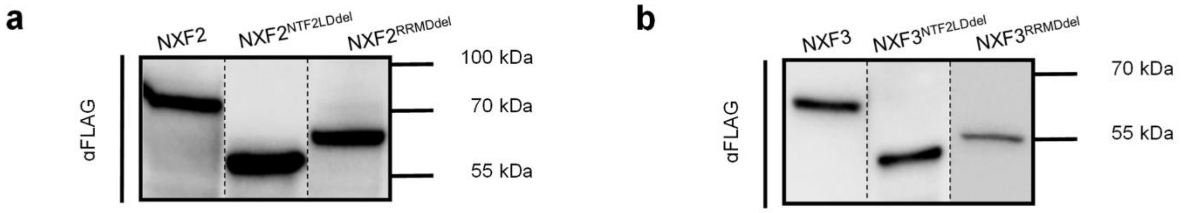


**Supplementary Figure 3. Western blot analysis of NXF2 and NXF3 proteins expressed from deletion constructs lacking the NTF2-like domain or RRM domain.**

a. The deletion of the NTF2-like domain and the RRM domain from the NXF2-FLAG expression construct results in the expression of truncated NXF2 proteins, when overexpressed in HEK cells, as visualized using a FLAG antibody in a Western blot. Calculated molecular weights are ~72 kDa for full-length NXF2-FLAG, ~55 kDa for $NXF2^{NTF2LDdel}$-FLAG, and ~63 kDa for $NXF2^{RRMDdel}$-FLAG. b. Western blot analysis of lysates derived from overexpression of NXF3 deletion constructs in HEK cells demonstrating expression of truncated FLAG-tagged NXF3 proteins compared to the wildtype. Calculated molecular weights are ~60 kDa for full-length NXF3-FLAG, ~43 kDa for $NXF3^{NTF2LDdel}$-FLAG, and ~51 kDa for $NXF3^{RRMDdel}$-FLAG. Uncropped images of all Western blots depicted are provided in the Source data file.

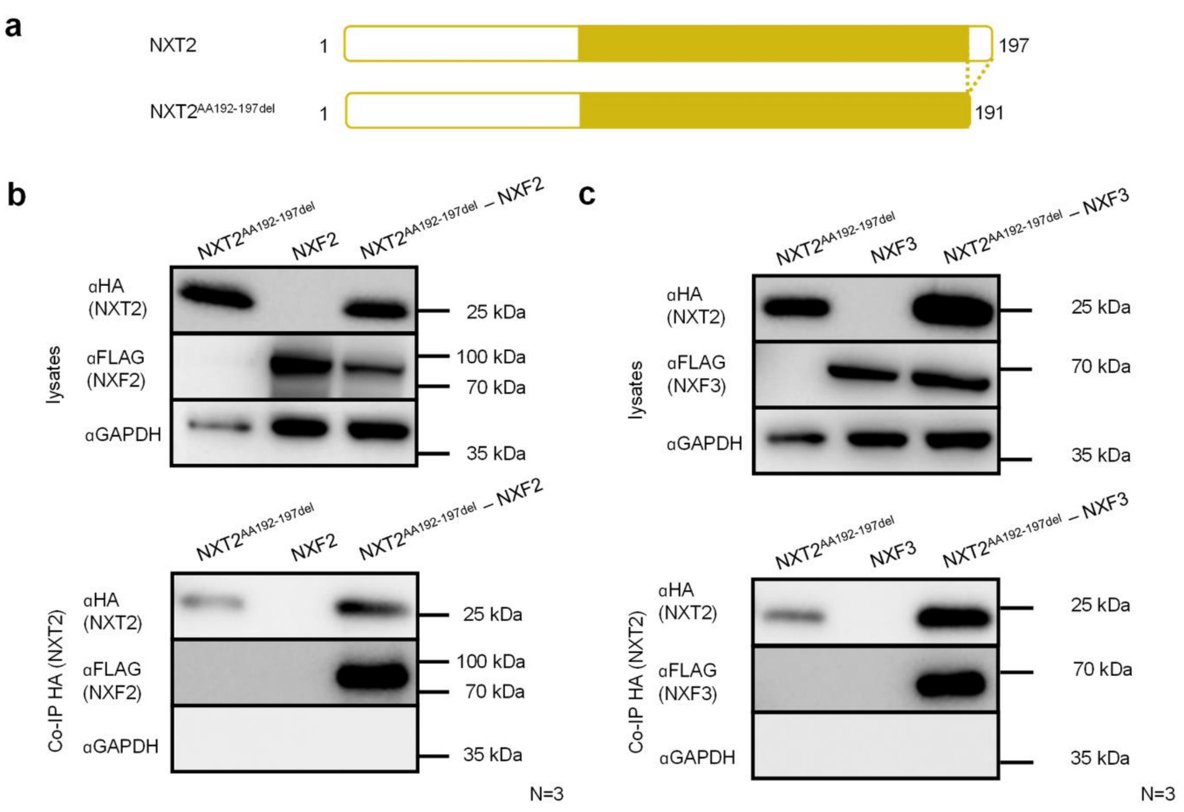


**Supplementary Figure 4. The NXT2 protein region downstream of the NTF2-like domain is not essential for binding to NXF2 and NXF3.**

a. Deletion of six C-terminal amino acids of NXT2-HA, located downstream of the predicted NTF2-like domain (amino acids 192-197 in NM_018698.5) truncates the protein. b. The C-terminal truncated NXT2-HA protein is still able to bind to NXF2-FLAG as demonstrated by Co-IP of protein lysates obtained from NXT2 and NXF2 constructs overexpressed in HEK293T cells and visualized in a Western blot. The top Western blot refers to protein lysates, while the bottom blot is derived from Co-IP. c. The interaction between NXT2$^{AA192-197del}$-HA and NXF3-FLAG persists even without the C-terminus of NXT2. Lysates are visualized at the top, Co-IP-samples at the bottom. Western blots demonstrate representative results of at least three replicates. Uncropped images of all Western blots depicted are provided in the Source data file.

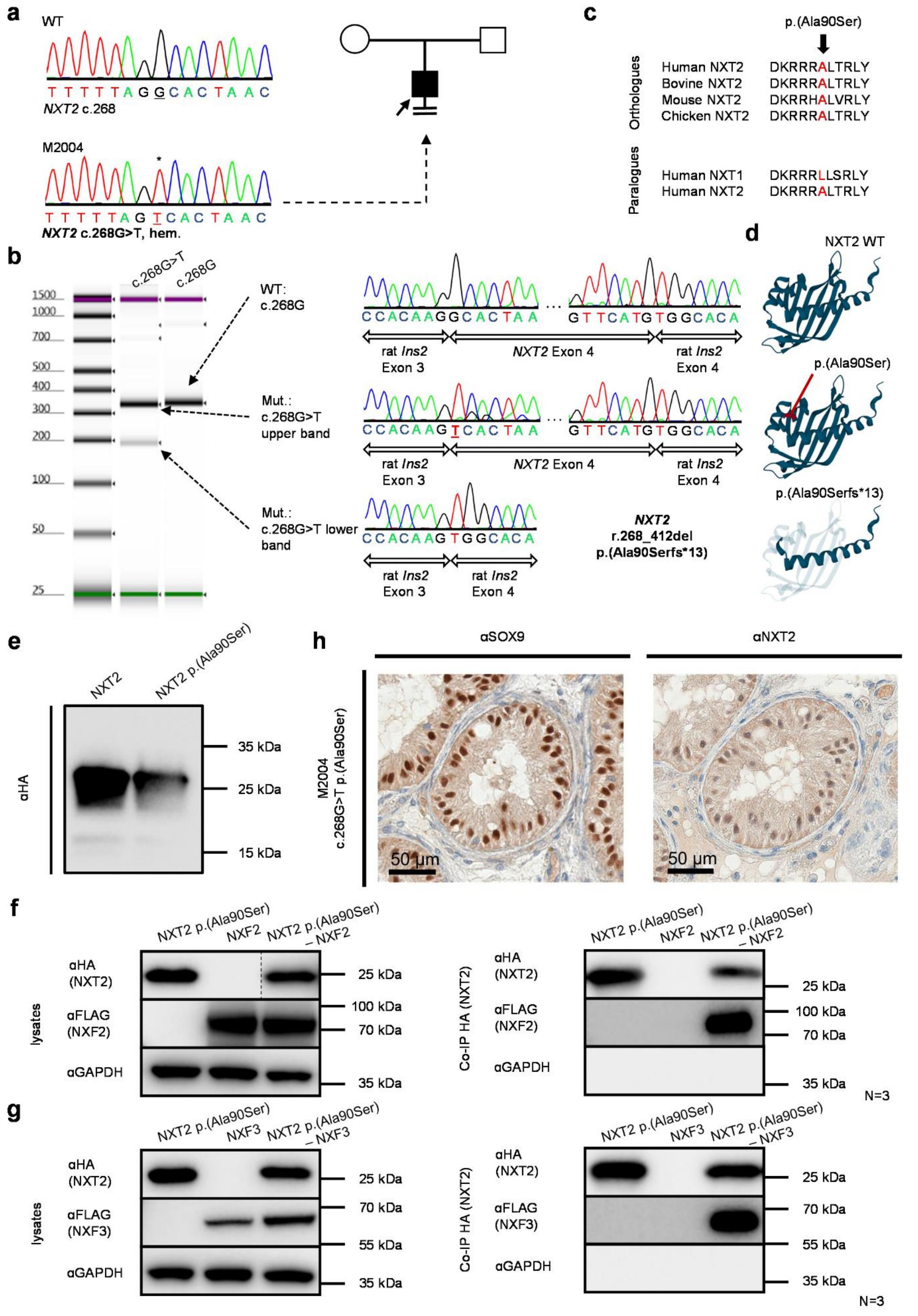
a
WT
T T T T T A G G C A C T A A C
NXT2 c.268
M2004
T T T T T A G T C A C T A A C
NXT2 c.268G>T, hem.
b
c.268G>T
c.268G
1500
1000
700
500
400
300
200
100
50
25
WT:
c.268G
Mut.:
c.268G>T
upper band
Mut.:
c.268G>T lower
band
C C A C A A G G C A C T A A
G T T C A T G T G G C A C A
rat Ins2
Exon 3
NXT2 Exon 4
rat Ins2
Exon 4
C C A C A A G T C A C T A A
C C A C A A G T G G C A C A
NXT2
r.268_412del
p.(Ala90Serfs*13)
c
p.(Ala90Ser)
Orthologues
Paralogues
Human NXT2 DKRRRALTRLY
Bovine NXT2 DKRRRALTRLY
Mouse NXT2 DKRRHALVRLY
Chicken NXT2 DKRRRALTRLY
Human NXT1 DKRRRLLSRLY
Human NXT2 DKRRRALTRLY
d
NXT2 WT
p.(Ala90Ser)
p.(Ala90Serfs*13)
e
NXT2
NXT2 p.(Ala90Ser)
αHA
35 kDa
25 kDa
15 kDa
h
αSOX9
αNXT2
M2004
c.268G>T p.(Ala90Ser)
50 µm
50 µm
f
NXT2 p.(Ala90Ser)
NXF2
NXT2 p.(Ala90Ser) – NXF2
lysates
αHA (NXT2)
αFLAG (NXF2)
αGAPDH
25 kDa
100 kDa
70 kDa
35 kDa
Co-IP HA (NXT2)
N=3
g
NXT2 p.(Ala90Ser)
NXF3
NXT2 p.(Ala90Ser) – NXF3
αHA (NXT2)
αFLAG (NXF3)
αGAPDH
25 kDa
70 kDa
55 kDa
35 kDa
N=3

**Supplementary Figure 5. The *NXT2* missense variant c.268G>T p.(Ala90Ser) in M2004 has an impact on splicing but no identified effect on the protein level.**

a. M2004 is positive for the hemizygous single nucleotide substitution c.268G>T in *NXT2* affecting the most 5’ nucleotide of exon four. DNA samples of other family members were not available for co-segregation analysis. b. In a minigene assay, the variant c.268G>T in *NXT2* leads to a skipping of exon 4 in a minor proportion of transcripts *in vitro*, resulting in a frameshift [r.268_412del p.(Ala90Serfs*13)]. c. The affected alanine residue is conserved in orthologs of NXT2 across bovine, mouse and chicken. In the paralog human protein NXT1, the amino acid is not preserved. d. AlphaFold2 predictions show wildtype, mutant (substituted residue = red) and truncated (loss of 107 amino acids caused by exon skipping) NXT2 3D structures. e, Western blot of overexpressed wildtype and mutant *NXT2* p.(Ala90Ser) demonstrated no effect of the amino acid substitution on protein stability. f, When overexpressed in HEK cells, NXT2 p.(Ala90Ser) and NXF2 are detectable (~26 kDa for NXT2 and ~72 kDa for NXF2) in a Western blot of cell lysates (left). The NXT2 p.(Ala90Ser) variant does not impair binding to NXF2 as shown by Co-IP experiments (right). g. Binding of NXT2 p.(Ala90Ser) to NXF3 is also not affected as shown by Co-IP. Western blots demonstrate representative results of at least three replicates. Uncropped images of all Western blots depicted are provided in the Source data file. h. Immunohistochemical staining of NXT2 in M2004 compared to control tissue demonstrated that NXT2 is still expressed in SOX9-positive Sertoli cells.

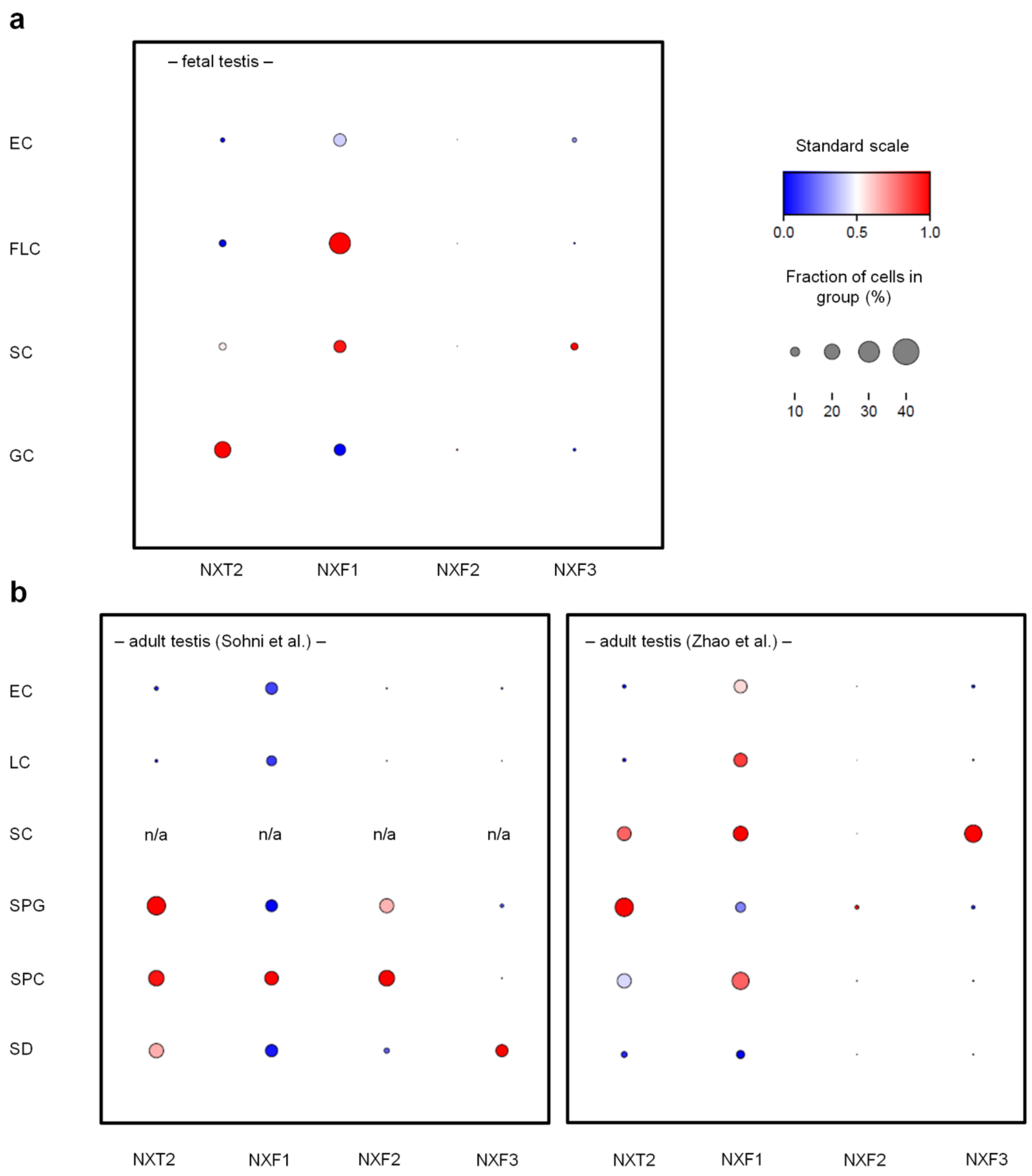


**Supplementary Figure. 6. scRNA-seq shows expression of nuclear export factor proteins in fetal male germ cells and adult testis.**

a. scRNA-seq dataset of fetal male germ cells[2] was analyzed via smart-DB[3]. *NXT2* reveals a strong expression in germ cells. *NXF1* is mainly expressed in fetal Leydig and in Sertoli cells. *NXF2* is generally weakly expressed and *NXF3* is expressed in fetal Sertoli but not in germ cells. b. In the adult testis, different scRNA-seq datasets[4,5] showed different expression patterns: Sohni et al. show *NXT2* expression mainly in spermatogonia, but also in spermatocytes and spermatids[4]. *NXF1* is ubiquitously expressed, *NXF2* is expressed in spermatogonia and spermatocytes and *NXF3* is mainly expressed in spermatids (left)[4]. In the dataset of Zhao et al. (right)[5] *NXT2*, *NXF1* and *NXF3* are also expressed in Sertoli cells (EC:

endothelial cells; FLC: fetal Leydig cells; LC: Leydig cells; SC: Sertoli cells; GC: germ cells; SPG: spermatogonia; SPC: spermatocytes; SD: spermatids).
