## Supplementary material for "NXT2 is the key player for nuclear RNA export in the human testis and critical for spermatogenesis": Source data

### Figure 1
### Source Data

g

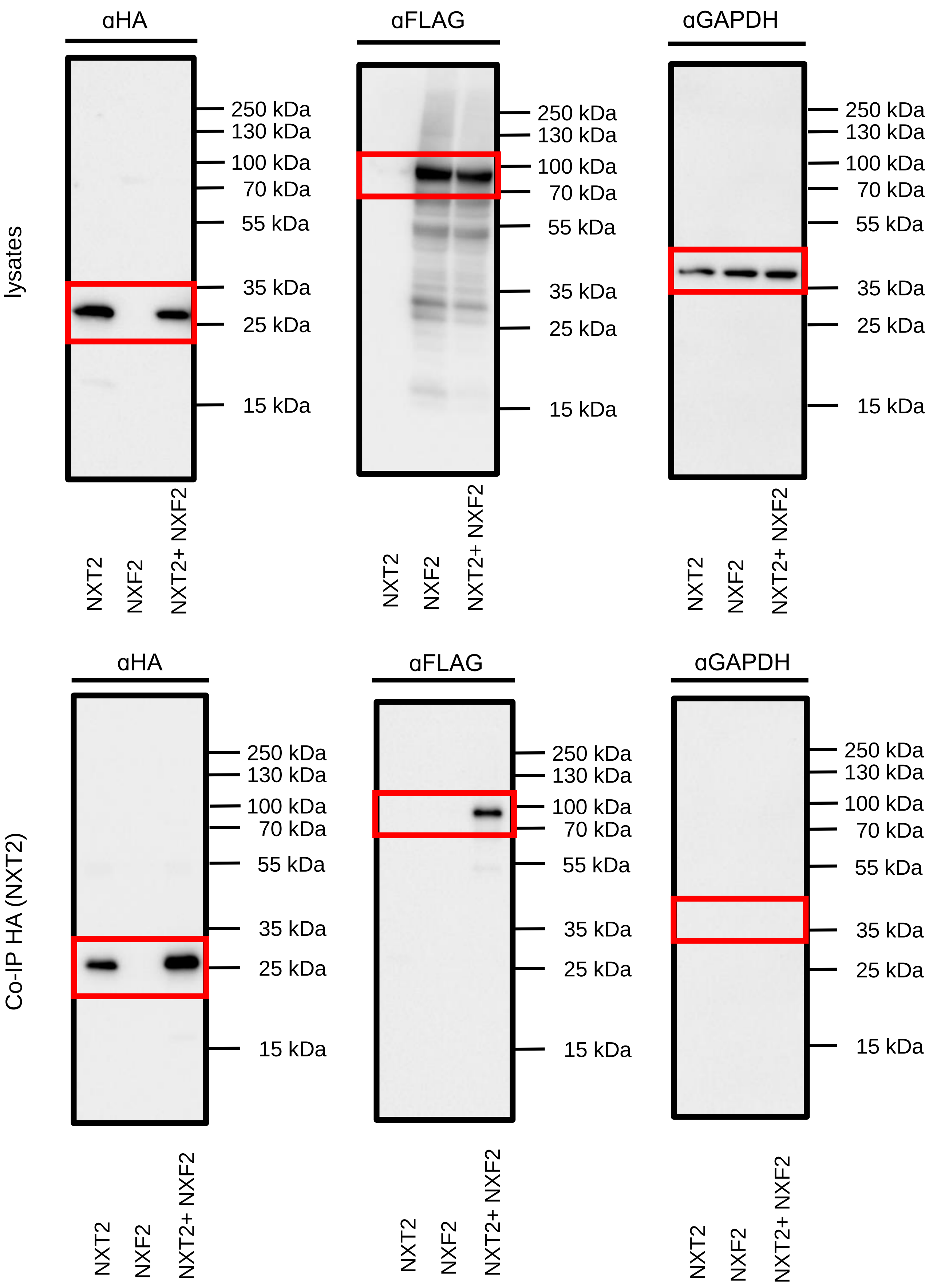

lysates
αHA
250 kDa
130 kDa
100 kDa
70 kDa
55 kDa
35 kDa
25 kDa
15 kDa
NXT2
NXF2
NXT2+ NXF2
αFLAG
αGAPDH
Co-IP HA (NXT2)

h

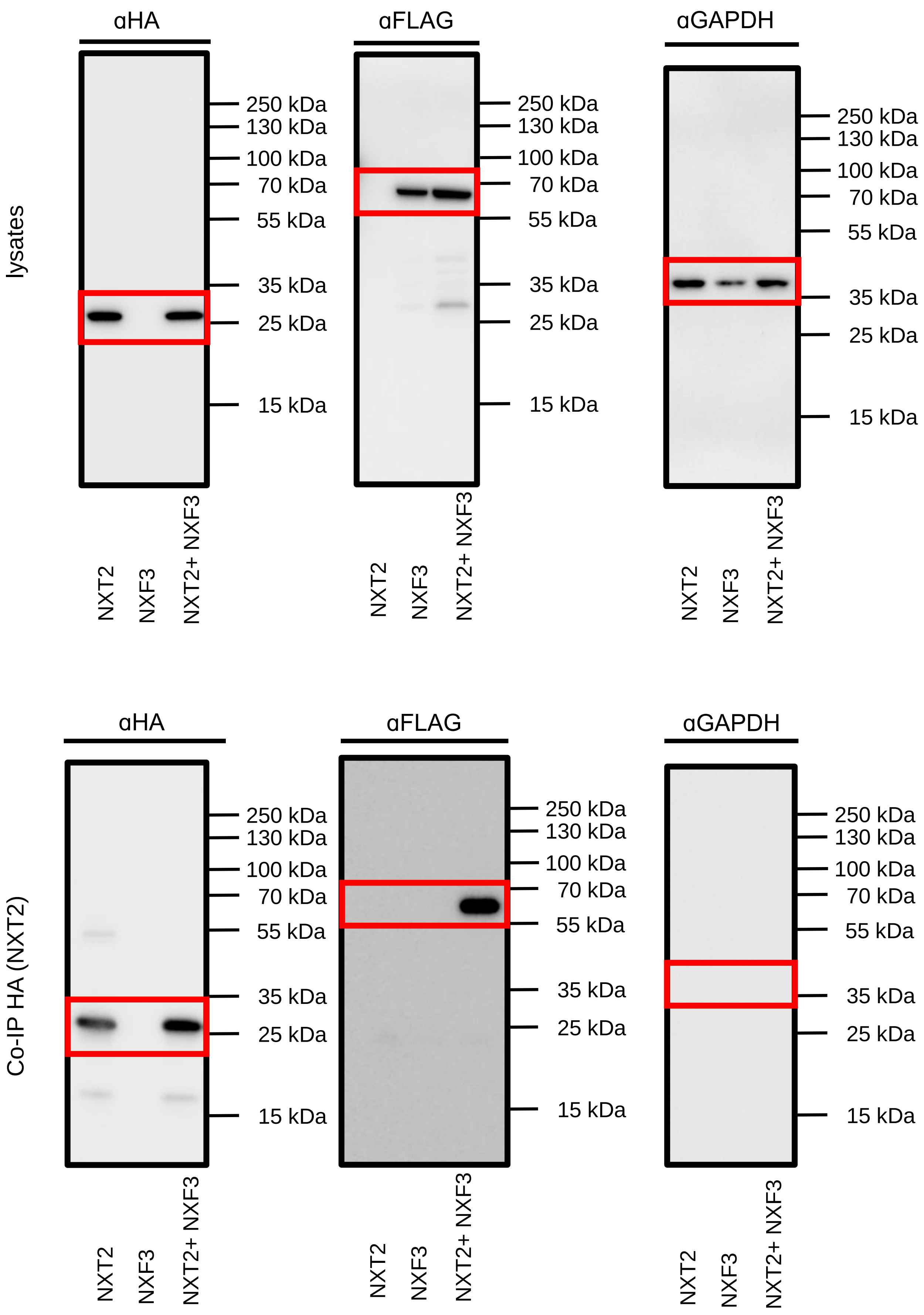
lysates
Co-IP HA (NXT2)
αHA
αFLAG
αGAPDH
250 kDa
130 kDa
100 kDa
70 kDa
55 kDa
35 kDa
25 kDa
15 kDa
NXT2
NXF3
NXT2+ NXF3

Figure 2
Source Data

b

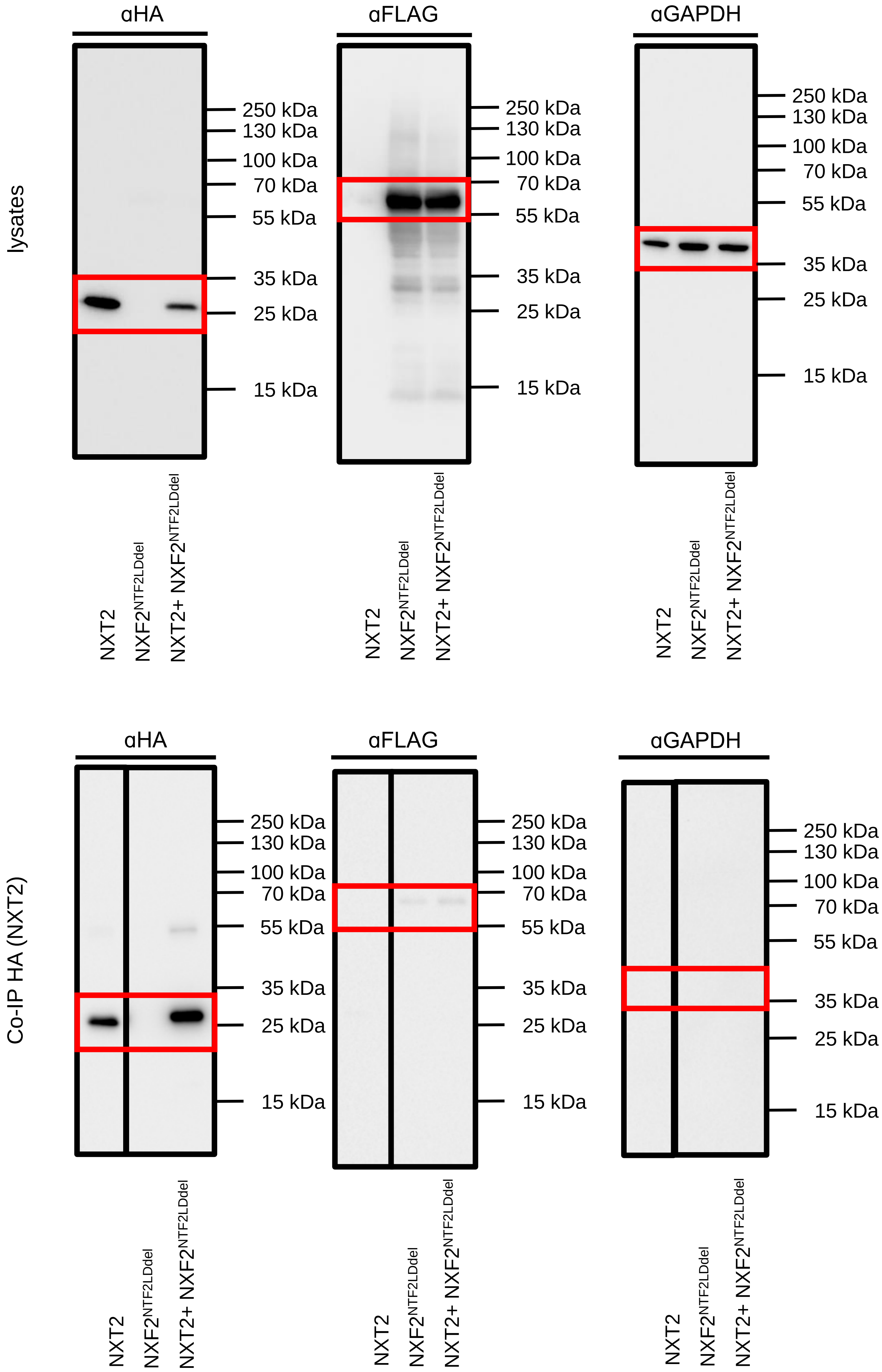

lysates
αHA
αFLAG
αGAPDH
250 kDa
130 kDa
100 kDa
70 kDa
55 kDa
35 kDa
25 kDa
15 kDa
NXT2
NXF2NTF2LDdel
NXT2+ NXF2NTF2LDdel
Co-IP HA (NXT2)

c

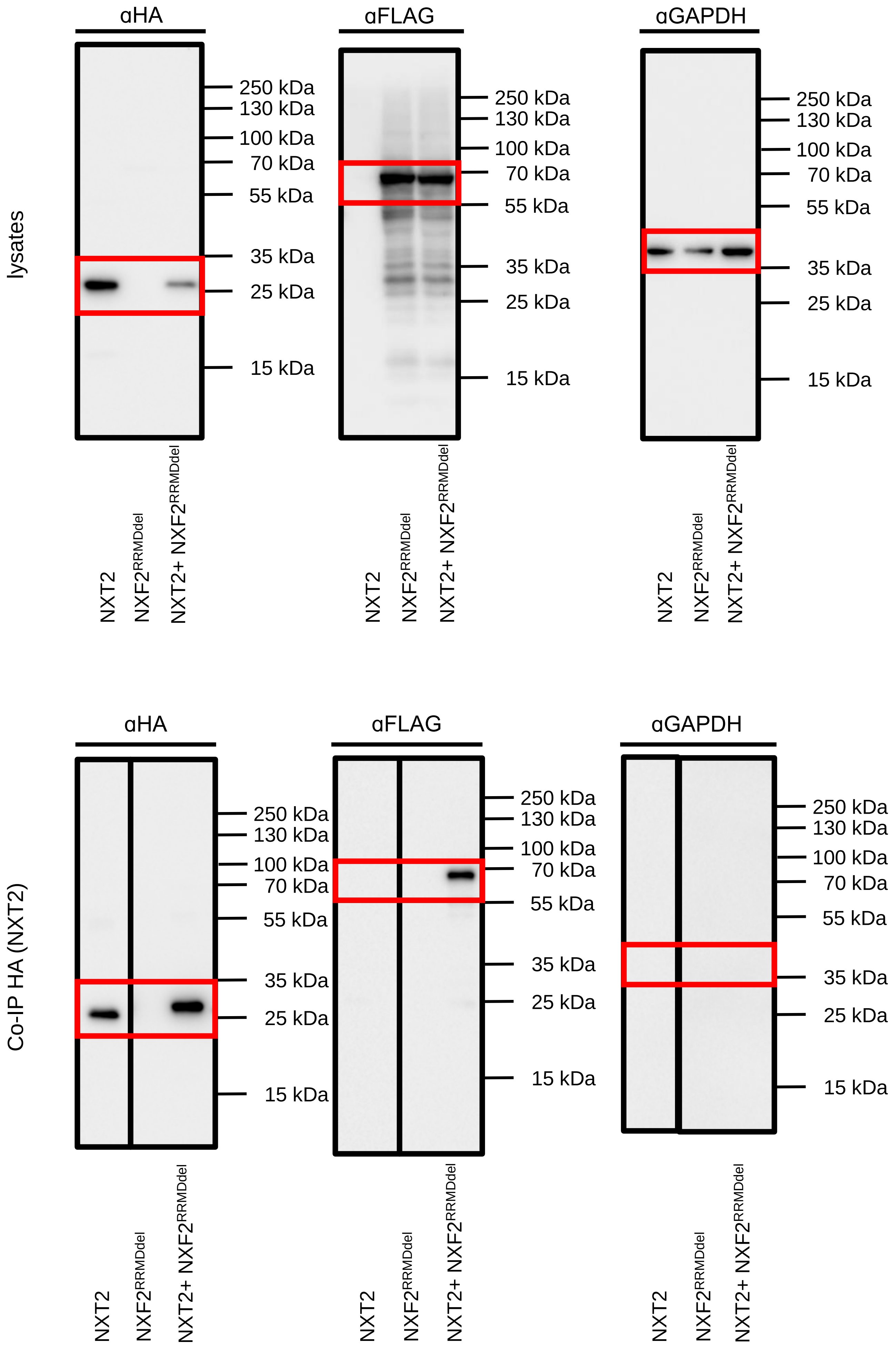
lysates
αHA
αFLAG
αGAPDH
250 kDa
130 kDa
100 kDa
70 kDa
55 kDa
35 kDa
25 kDa
15 kDa
NXT2
NXF2RRMDdel
NXT2+ NXF2RRMDdel
Co-IP HA (NXT2)

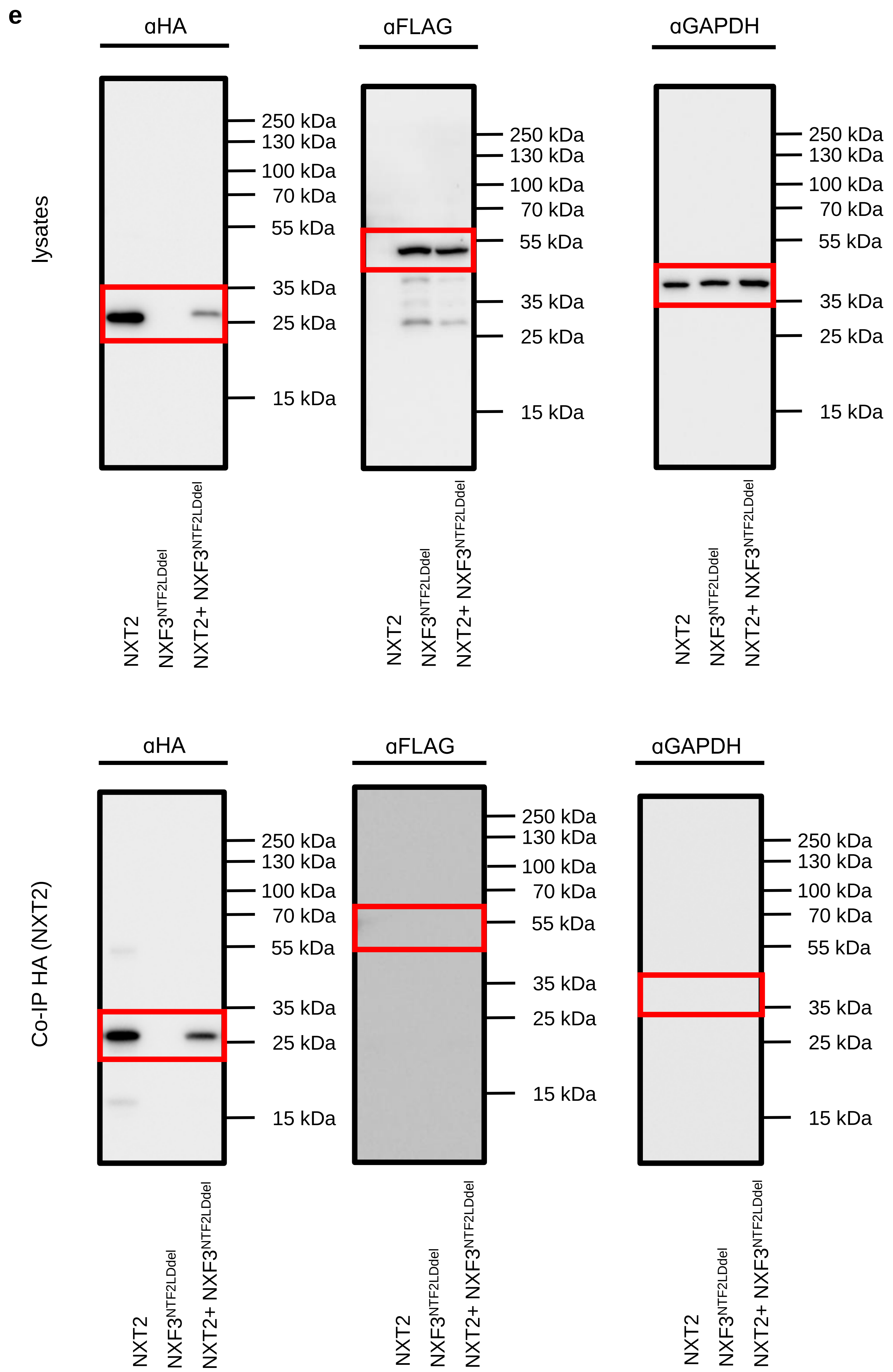
e
lysates
Co-IP HA (NXT2)
αHA
αFLAG
αGAPDH
250 kDa
130 kDa
100 kDa
70 kDa
55 kDa
35 kDa
25 kDa
15 kDa
NXT2
NXF3NTF2LDdel
NXT2+ NXF3NTF2LDdel

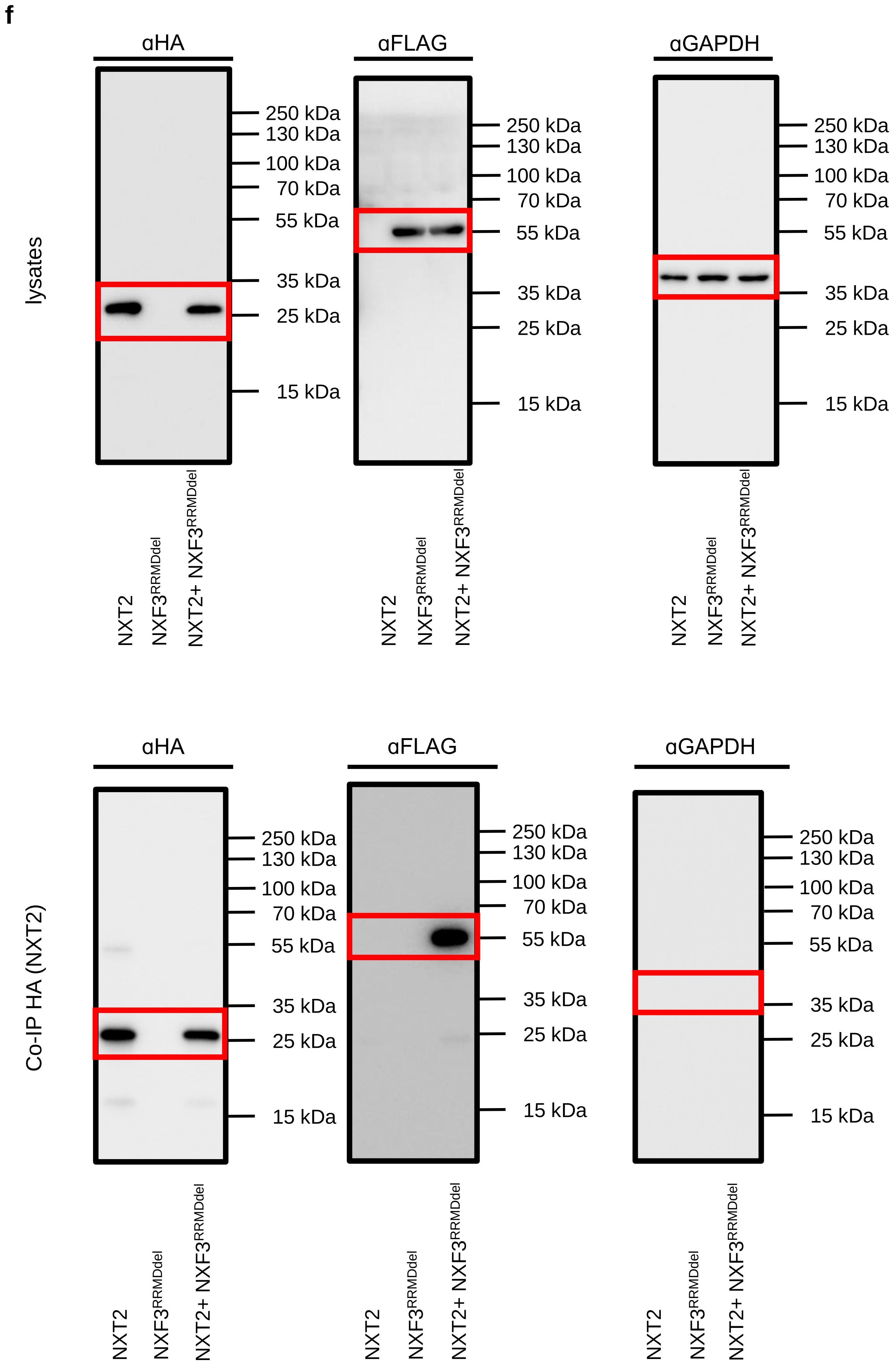

f
lysates
αHA
αFLAG
αGAPDH
250 kDa
130 kDa
100 kDa
70 kDa
55 kDa
35 kDa
25 kDa
15 kDa
NXT2
NXF3^RRMDdel
NXT2+ NXF3^RRMDdel
Co-IP HA (NXT2)

### Figure 4
### Source Data

c

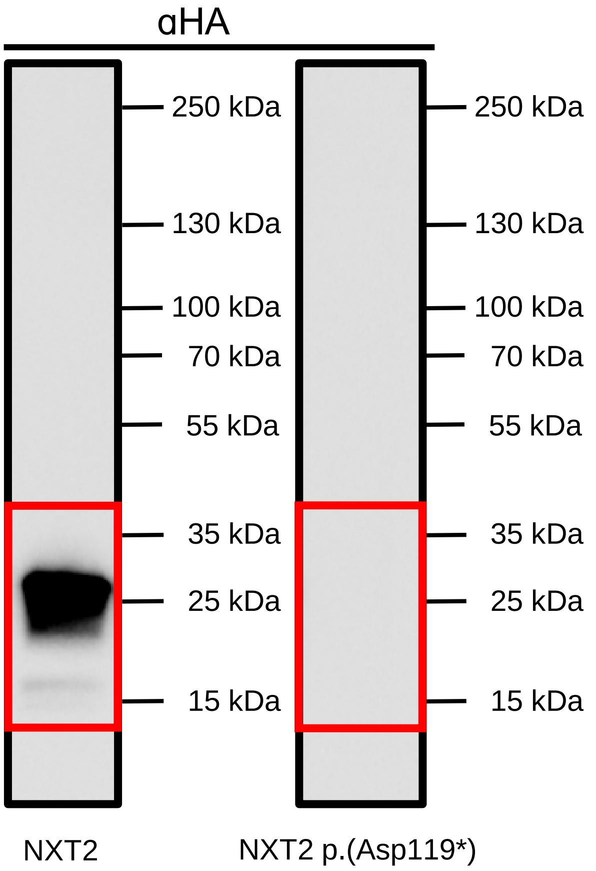
αHA
250 kDa
130 kDa
100 kDa
70 kDa
55 kDa
35 kDa
25 kDa
15 kDa
250 kDa
130 kDa
100 kDa
70 kDa
55 kDa
35 kDa
25 kDa
15 kDa
NXT2
NXT2 p.(Asp119*)

Figure 6
Source Data

d

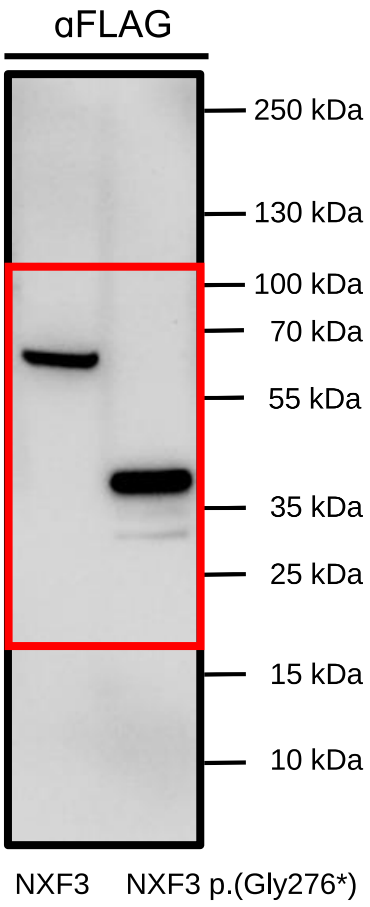
αFLAG
250 kDa
130 kDa
100 kDa
70 kDa
55 kDa
35 kDa
25 kDa
15 kDa
10 kDa
NXF3
NXF3 p.(Gly276*)

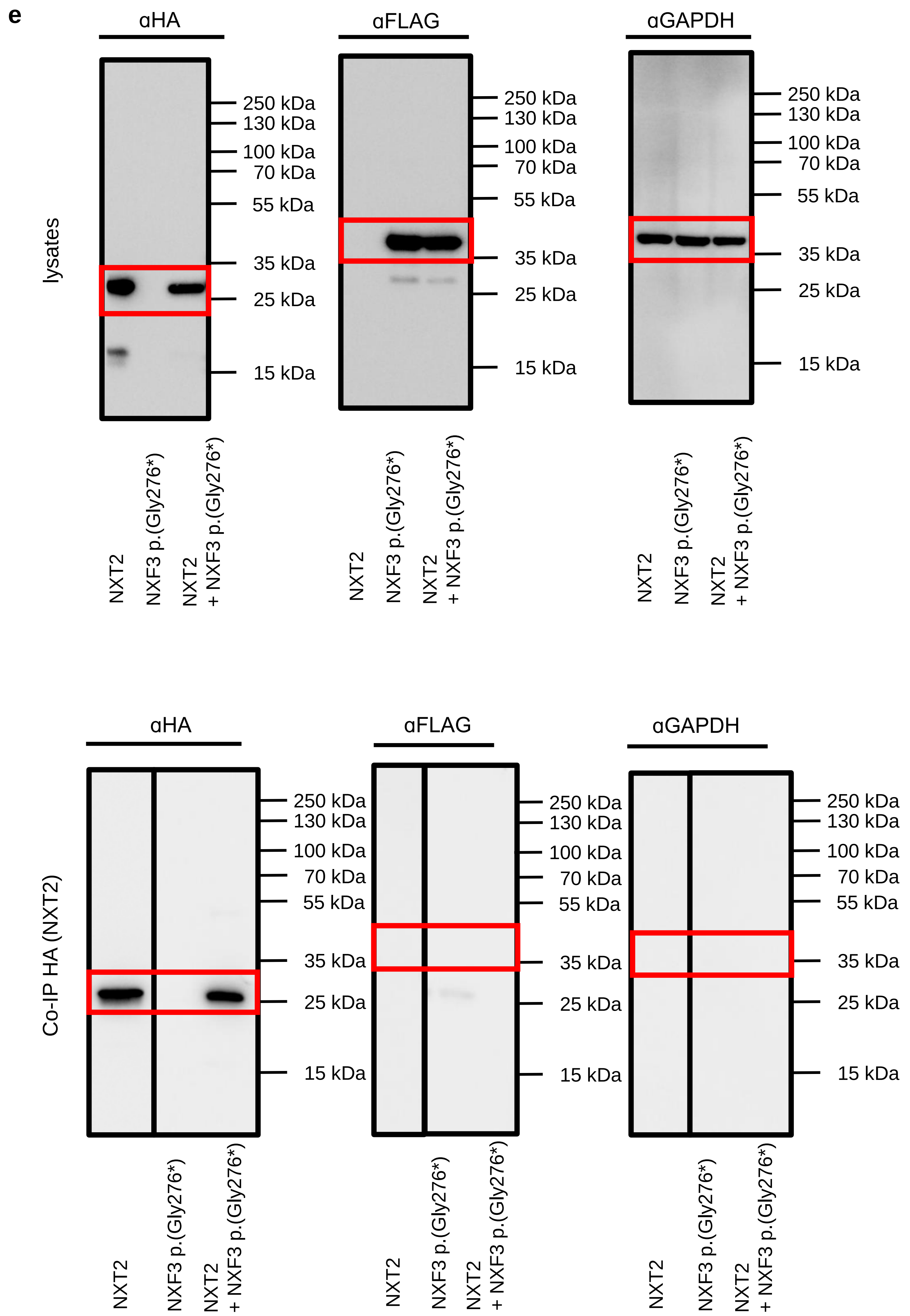
e
αHA
αFLAG
αGAPDH
lysates
250 kDa
130 kDa
100 kDa
70 kDa
55 kDa
35 kDa
25 kDa
15 kDa
NXT2
NXF3 p.(Gly276*)
NXT2
+ NXF3 p.(Gly276*)
Co-IP HA (NXT2)

Supplementary
Figure 3
Source Data

a

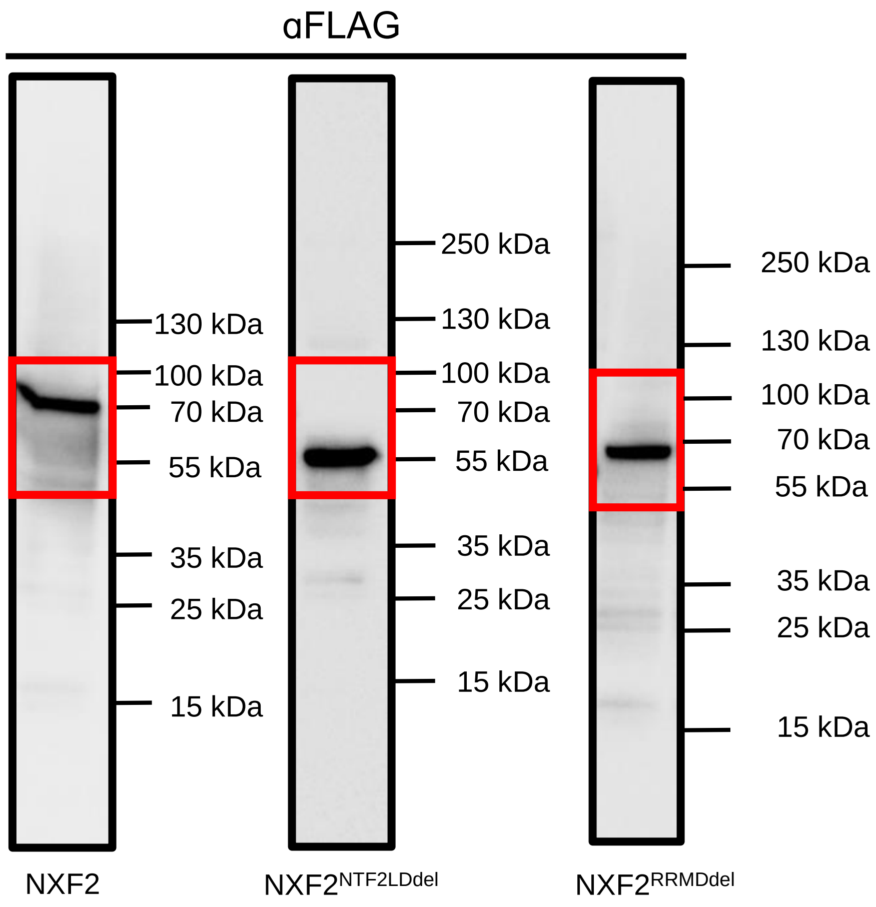
αFLAG
130 kDa
100 kDa
70 kDa
55 kDa
35 kDa
25 kDa
15 kDa
250 kDa
130 kDa
100 kDa
70 kDa
55 kDa
35 kDa
25 kDa
15 kDa
250 kDa
130 kDa
100 kDa
70 kDa
55 kDa
35 kDa
25 kDa
15 kDa
NXF2
NXF2NTF2LDdel
NXF2RRMDdel

**b**

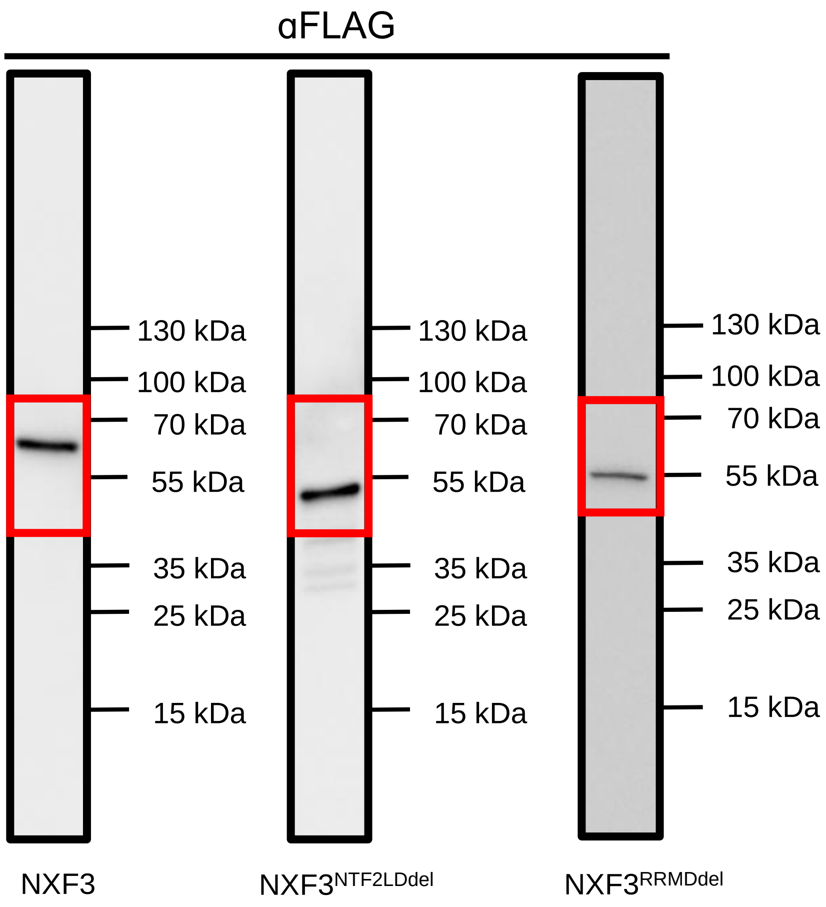

αFLAG
130 kDa
100 kDa
70 kDa
55 kDa
35 kDa
25 kDa
15 kDa
130 kDa
100 kDa
70 kDa
55 kDa
35 kDa
25 kDa
15 kDa
130 kDa
100 kDa
70 kDa
55 kDa
35 kDa
25 kDa
15 kDa
NXF3
NXF3NTF2LDdel
NXF3RRMDdel

### Supplementary Figure 4 Source Data

**b**

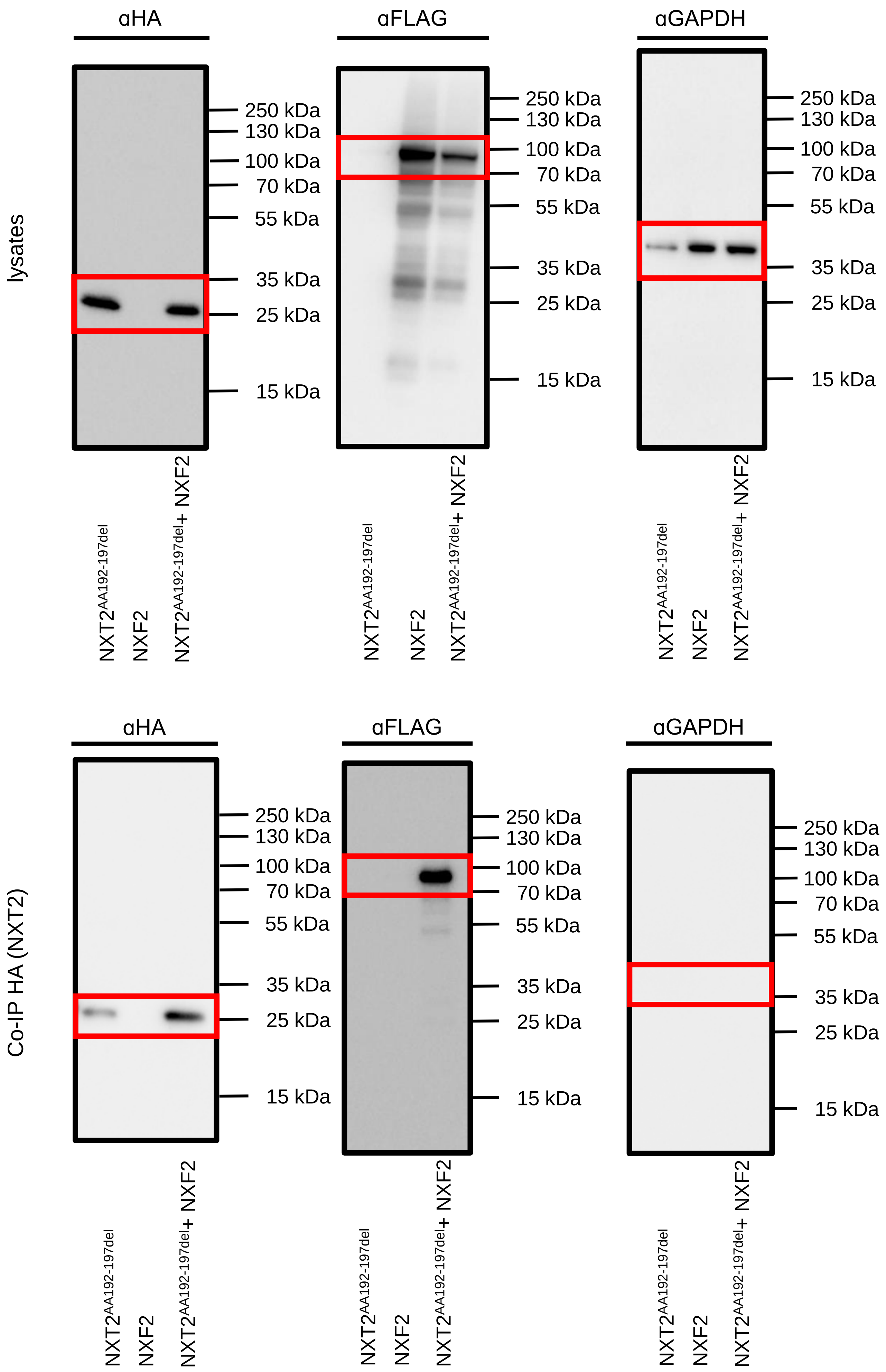

αHA
αFLAG
αGAPDH
lysates
250 kDa
130 kDa
100 kDa
70 kDa
55 kDa
35 kDa
25 kDa
15 kDa
NXT2AA192-197del
NXF2
NXT2AA192-197del+ NXF2
Co-IP HA (NXT2)

c

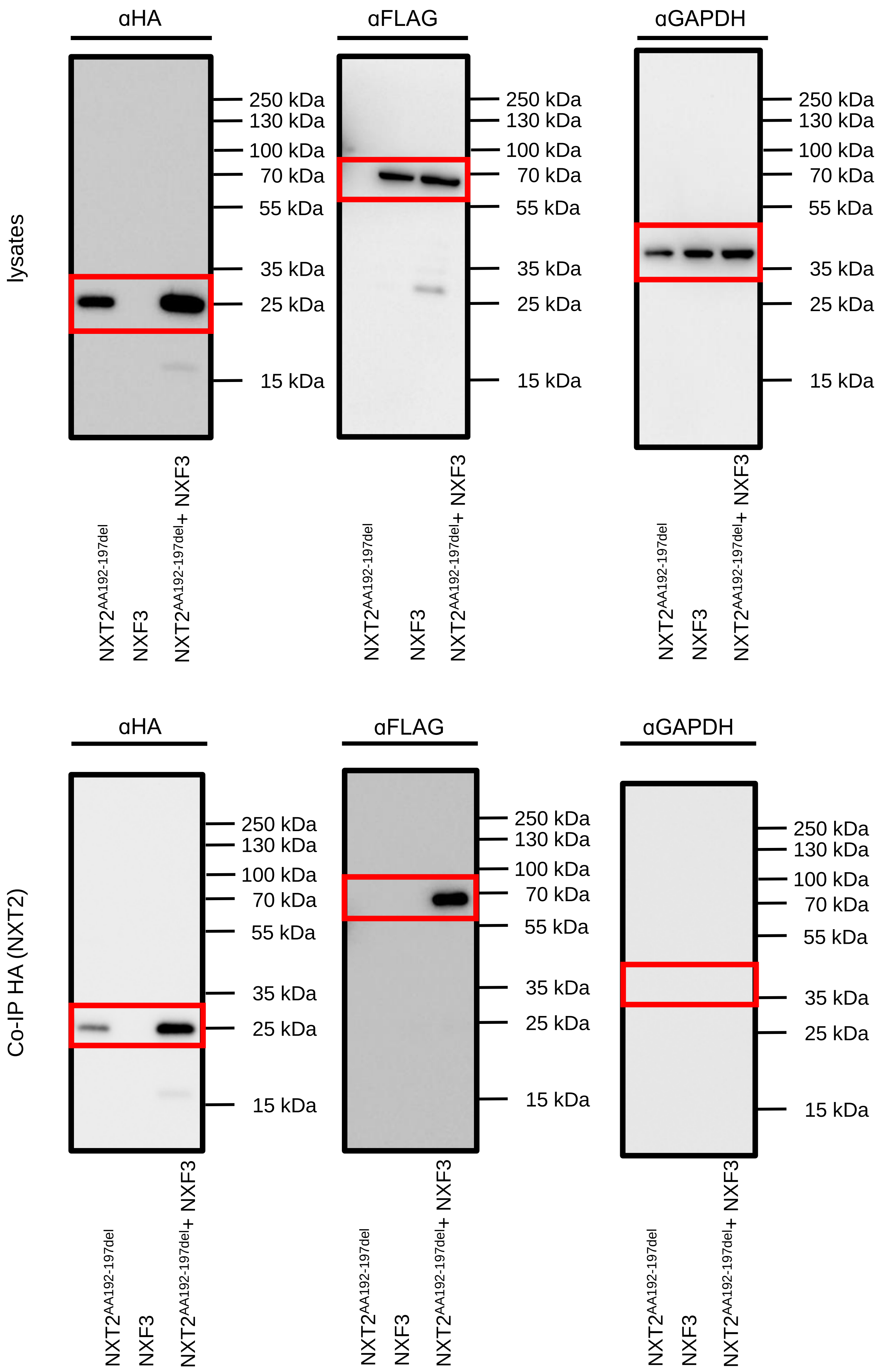
lysates
αHA
αFLAG
αGAPDH
250 kDa
130 kDa
100 kDa
70 kDa
55 kDa
35 kDa
25 kDa
15 kDa
$NXT2^{AA192-197del}$
NXF3
$NXT2^{AA192-197del}$+ NXF3
Co-IP HA (NXT2)
αHA
αFLAG
αGAPDH
$NXT2^{AA192-197del}$
NXF3
$NXT2^{AA192-197del}$+ NXF3

### Supplementary Figure 5 Source Data

e

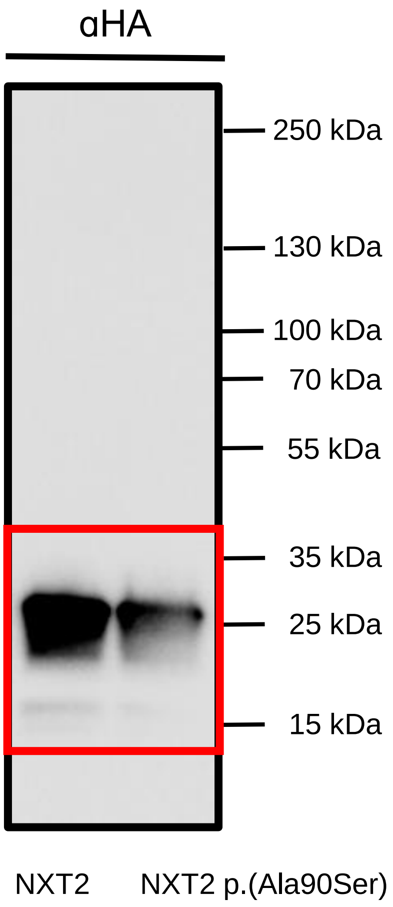
αHA
250 kDa
130 kDa
100 kDa
70 kDa
55 kDa
35 kDa
25 kDa
15 kDa
NXT2
NXT2 p.(Ala90Ser)

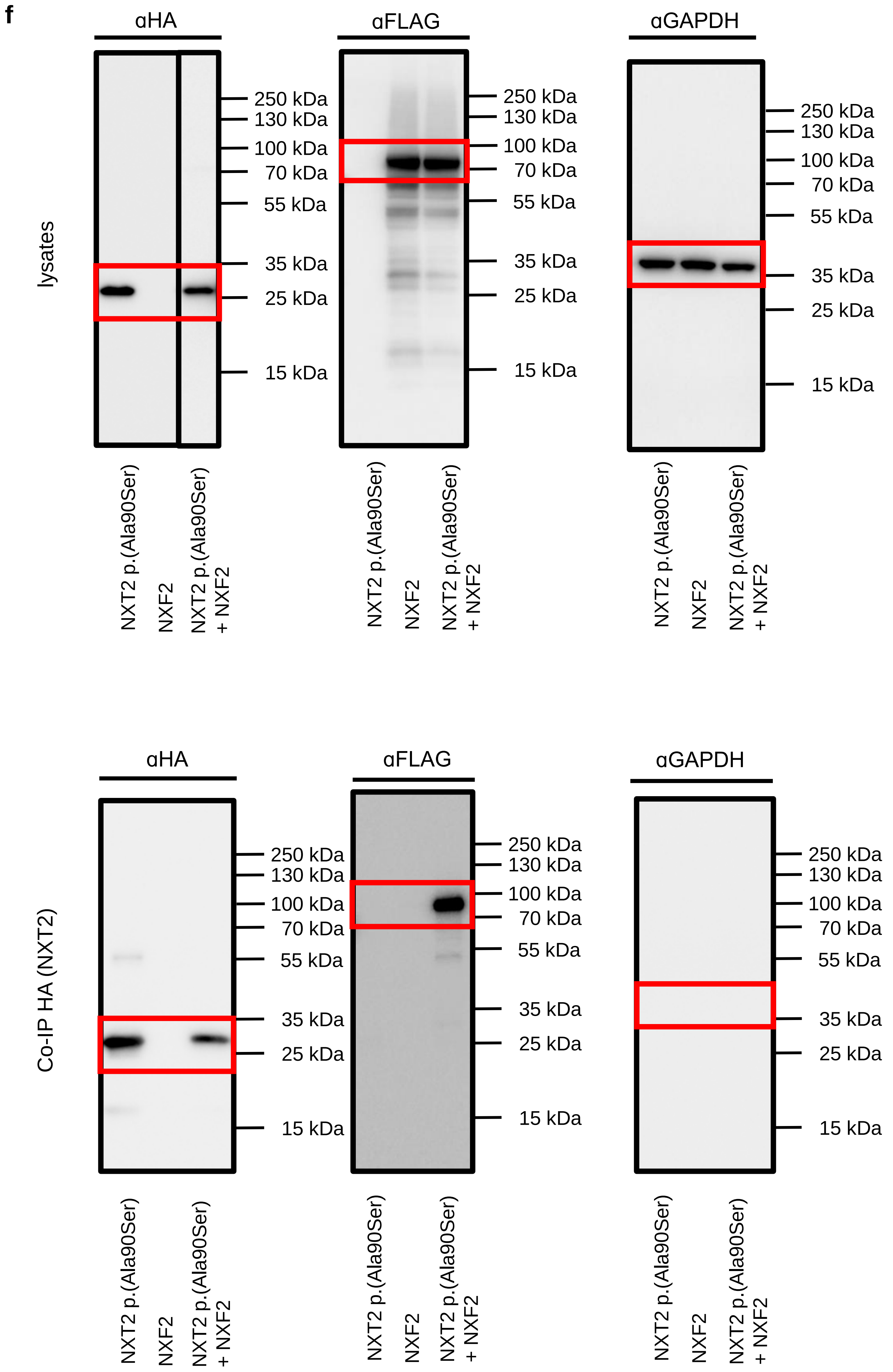
f
lysates
αHA
αFLAG
αGAPDH
250 kDa
130 kDa
100 kDa
70 kDa
55 kDa
35 kDa
25 kDa
15 kDa
NXT2 p.(Ala90Ser)
NXF2
NXT2 p.(Ala90Ser) + NXF2
Co-IP HA (NXT2)
αHA
αFLAG
αGAPDH
250 kDa
130 kDa
100 kDa
70 kDa
55 kDa
35 kDa
25 kDa
15 kDa
NXT2 p.(Ala90Ser)
NXF2
NXT2 p.(Ala90Ser) + NXF2

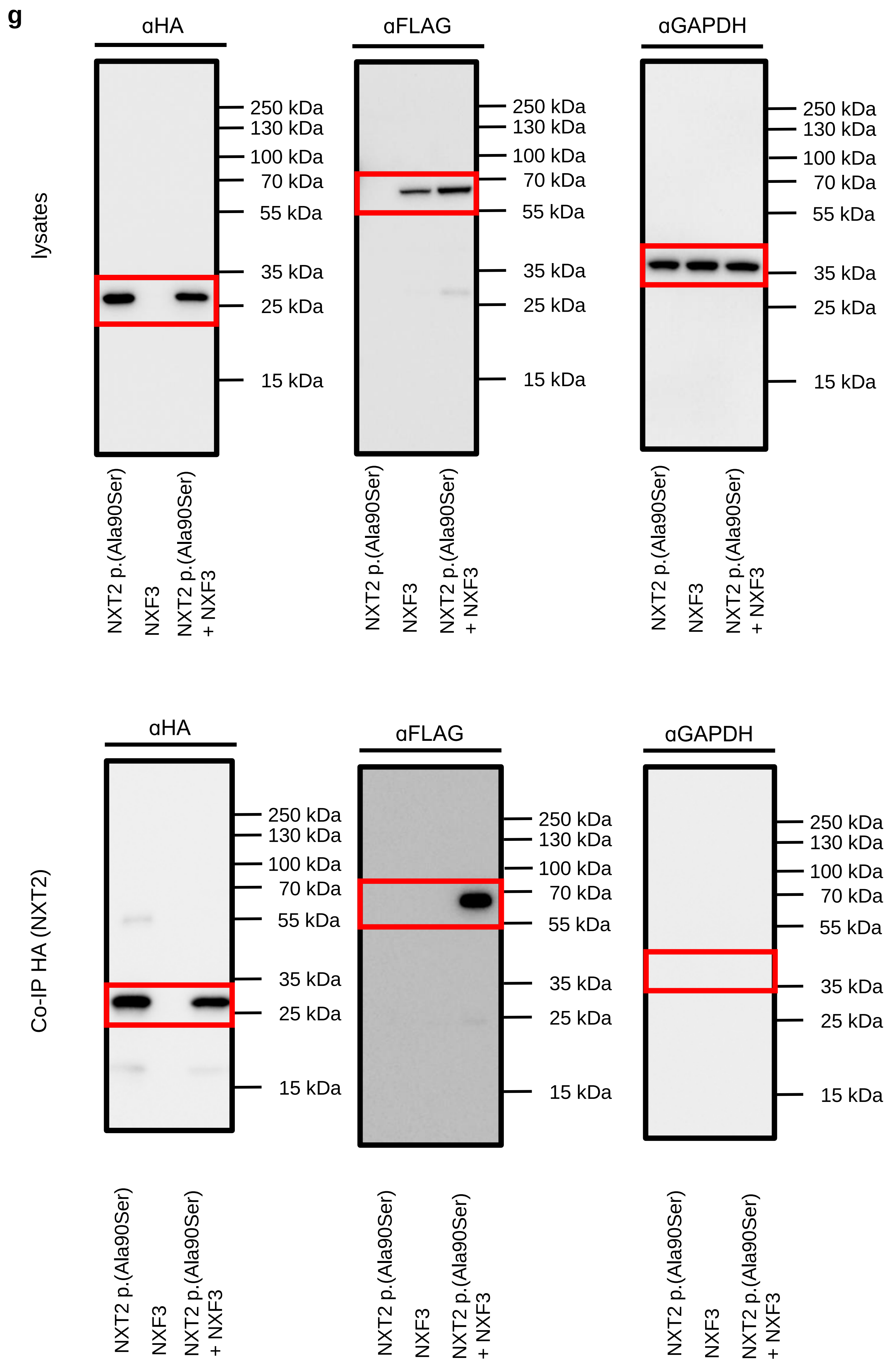
g
lysates
αHA
αFLAG
αGAPDH
250 kDa
130 kDa
100 kDa
70 kDa
55 kDa
35 kDa
25 kDa
15 kDa
NXT2 p.(Ala90Ser)
NXF3
NXT2 p.(Ala90Ser) + NXF3
Co-IP HA (NXT2)
αHA
αFLAG
αGAPDH
250 kDa
130 kDa
100 kDa
70 kDa
55 kDa
35 kDa
25 kDa
15 kDa
NXT2 p.(Ala90Ser)
NXF3
NXT2 p.(Ala90Ser) + NXF3
